# Cadherin-26 drives macrophage alternative activation via suppressing STUB1-mediated IL-4Rα ubiquitination in asthma

**DOI:** 10.1101/2024.08.01.24311333

**Authors:** Gongqi Chen, Shengchong Chen, Chunli Huang, Wei Gu, Huiru Jie, Lu Zhao, Weiqiang Kong, Jiali Gao, Yuchen Feng, Lingling Yi, Peisong Gao, Guohua Zhen

## Abstract

**Introduction:** IL-4 receptor (IL-4R)-mediated alternative activation of macrophage drives type 2 airway inflammation. Cadherin-26 (CDH26) upregulates epithelial type II IL-4R signaling in asthma. However, whether CDH26 contributes to type I IL-4R-mediated macrophage activation and the mechanism by which CDH26 upregulates IL-4R expression remains unknown.

**Methods:** CDH26 expression in bronchoalveolar lavage cells of asthma patients was examined using quantitative PCR and immunostaining. Airway inflammation and macrophage activation were assessed in ovalbumin-sensitized and challenged *Cdh26^fl/fl^Lyz2Cre* and control mice. Mechanistic experiments included IL-4Rα degradation and ubiquitination assay, CDH26 co-immunoprecipitation and mass spectrometry analysis. *Cdh26* siRNA encapsulated lipid nanoparticles were used to treat the mouse model.

**Results:** CDH26 expression was enhanced in bronchoalveolar lavage cells from patients with eosinophilic asthma and was localized to macrophages. Airway eosinophilia, mucous cell metaplasia and macrophage alternative activation were significantly suppressed in ovalbumin-challenged *Cdh26^fl/fl^Lyz2Cre* mice compared to control mice. *Cdh26* deficiency inhibited the expression of M2 markers as well as IL-4Rα expression in mouse bronchoalveolar lavage macrophages, cultured bone marrow-derived macrophages, and primary lung macrophages. Furthermore, CDH26 knockdown enhanced whereas CDH26 overexpression suppressed IL-4Rα ubiquitination and proteasomal degradation in vitro. Mechanistically, CDH26 directly interacts with STUB1 and suppresses the binding of STUB1 to IL-4Rα and subsequent ubiquitination-proteasomal degradation. *Cdh26* siRNA encapsulated lipid nanoparticles markedly alleviated airway eosinophilia, mucus metaplasia and macrophage alternative activation in the mouse model.

**Conclusions:** CDH26 interacts with STUB1 and suppresses STUB1-mediated IL-4Rα ubiquitination-proteasomal degradation, thereby amplifying IL-4R signaling in macrophages in asthma. CDH26 is a potential therapeutic target for asthma.

**Take-Home Message:** Macrophage CDH26 is upregulated in asthma patients, and *Cdh26* deficiency suppresses macrophage alternative activation. CDH26 directly interacts with STUB1 and inhibits STUB1-mediated IL-4Rα ubiquitination and degradation, thus amplifying IL-4R signaling.

## Introduction

Asthma is a chronic airway disease that characterized by airway inflammation, mucus overproduction, airway remodeling, and airway hyperresponsiveness (1). Asthma affects approximately 358 million people worldwide and is among the top five respiratory diseases posing a threat to global health (2). Severe asthma imposes significant burdens on individuals and society, with the potential for fatal outcomes (3). Eosinophilic airway inflammation driven by type 2 immune response is commonly observed in severe asthma patients (4).

Macrophages play an important role in environmental allergen-induced airway inflammation in asthma (5). Macrophages exhibit different activation states in response to various stimuli (6). When activated by microbes-induced factors such as interferon-γ and reactive oxygen species, macrophages switch to a pro-inflammatory state named as classical activation (M1 macrophages) and eliminate engulfed microbes (7). On the other hand, when activated by allergen-induced type 2 cytokines including IL-4 and IL-13, macrophages are alternatively activated (M2 macrophages) to promote tissue repair, remodeling, and wound healing (8). Macrophage alternative activation is involved in the development of allergic diseases including asthma (9). Alternatively activated macrophages release high levels of IL-13 and chemokines including CCL17, CCL18, CCL22, and CCL24, which activate Th2 cells and promote eosinophilic inflammation (10).

The IL-4R-STAT6 signaling pathway plays a crucial role in the alternative activation of macrophages (11). There are two types of IL-4R (12). Type I IL-4R consists of IL-4Rα and IL-2Rγ (γc) subunits and is expressed in hematopoietic cells including lymphocytes and macrophages (13). Type II IL-4R, composed of IL-4Rα and IL-13Rα1 subunits, is present in hematopoietic and non-hematopoietic cells including airway epithelial cells and smooth muscle cells (14). Type I IL-4R specifically binds to IL-4, leading to downstream JAK1-STAT6 activation and macrophages alternative activation (15). Meanwhile, the occupied IL-4R is degraded to terminate and prevent sustained activation of IL-4R signaling. There are several mechanisms underlying IL-4R degradation including endocytosis and ubiquitin-proteasome system(16–18). Wei and colleagues demonstrated that STIP1 homology and U-box-containing protein 1 (STUB1), an E3 ligase, interacts with IL-4Rα, a subunit shared by type I and type II IL-4R, to lead to its ubiquitination and proteasomal degradation (17).

Cadherin family members including cadherin-1 play important roles in maintaining the adhesive connections between airway epithelial cells as well as regulating epithelial cell proliferation and differentiation (19). CDH26, a non-classical member of cadherin family, contributes to apicobasal polarization of airway epithelial cells (20). CDH26 has long been associated to type 2 inflammation-related diseases including asthma, helminth infection, eosinophilic gastritis, and esophagitis (21–24), but the precise role of CDH26 in these diseases is unclear. We recently reported that CDH26 promotes airway eosinophilia and mucus overproduction via upregulating epithelial type II IL-4R signaling (25). However, whether CDH26 contributes to type I IL-4R-mediated macrophage activation and the mechanism by which CDH26 upregulates IL-4R expression remains unknown.

In this study, we observed elevated CDH26 expression in lung macrophages from patients with eosinophilic asthma. Macrophage-specific *Cdh26* deficiency significantly suppressed macrophage alternative activation, airway eosinophilia, and mucus overproduction in a mouse model of allergic airway inflammation. CDH26 knockdown or overexpression accelerated or inhibited IL-4Rα proteasomal degradation in vitro, respectively. Mechanistically, CDH26 interacts with STUB1 to suppress its binding to IL-4Rα, thus inhibiting IL-4Rα ubiquitination and degradation. Intriguingly, *Cdh26* siRNA encapsulated lipid nanoparticles effectively alleviated airway eosinophilia and mucus overproduction in the mouse model.

## Materials and methods

For additional information on the materials and methods used in this study, please refer to the online supplementary materials.

### Subjects

We recruited 17 control subjects, 25 eosinophilic asthma patients (sputum eosinophils > 3%), and 7 non-eosinophilic asthma patients (sputum eosinophil ≤ 3%) (26). None of the participants had a history of smoking or received inhaled or oral corticosteroids or leukotriene antagonists. All participants provided written informed consent, and the study was approved by the ethics committee of Tongji Hospital, Tongji Medical College, Huazhong University of Science and Technology.

### Murine model of allergic airway inflammation

*Cdh26^fl/fl^Lyz2Cre* mice on a C57BL/6N background were generated and provided by GemPharmatech (Nanjing, China). Exon 4 of *Cdh26* gene was deleted in *Cdh26^fl/fl^Lyz2Cre* mice. All experimental procedures were approved by the Animal Care and Use Committee of Tongji Hospital, Tongji Medical College, Huazhong University of Science and Technology.

### Statistical analysis

Data analysis was performed using Prism version 8 (GraphPad Software). For normally distributed data, we calculated the means ± standard deviation (SD) and performed Student’s *t* test or one-way ANOVA followed by Tukey’s multiple comparison test to compare across groups. For non-normally distributed data, we calculated medians with interquartile ranges and performed Mann-Whitney test. Spearman’s correlation coefficient was used to assess correlation. Statistical significance was defined as *p* < 0.05.

## Results

### 1. CDH26 expression is upregulated in BAL macrophages from eosinophilic asthma patients

Subject characteristics are summarized in Table 1. We initially examined the expression of CDH26 in BAL cells from our cohort of mild-moderate asthma (n=32) and control subjects (n=17) using quantitative PCR. *CDH26* transcript levels were significantly higher in BAL cells from asthma patients compared to control subjects (Figure 1A). We further classified asthma patients as eosinophilic asthma (n=25, sputum eosinophils > 3%) and non-eosinophilic asthma (n=7, sputum eosinophils ≤ 3%) (26). *CDH26* transcript levels were significantly higher in BAL cells from eosinophilic asthma patients compared to control subjects, but no significant difference was observed between non-eosinophilic asthma patients and control subjects (Figure 1B). Moreover, *CDH26* transcript levels in BAL cells of asthma patients are positively correlated with fractional exhaled nitric oxide (FeNO), sputum eosinophil percentages, and serum IgE (Figures 1C-D, S1E). Additionally, *CDH26* transcript levels and markers for macrophage alternative activation including *CD206*, *FIZZ1* and *CCL17* were all significantly enhanced in sputum cells from asthma patients compared to control subjects (Figures S1A-D), and *CDH26* and *CCL17* expression are correlated with each other in asthma patients (Figures S1F-G).

**Table 1.**
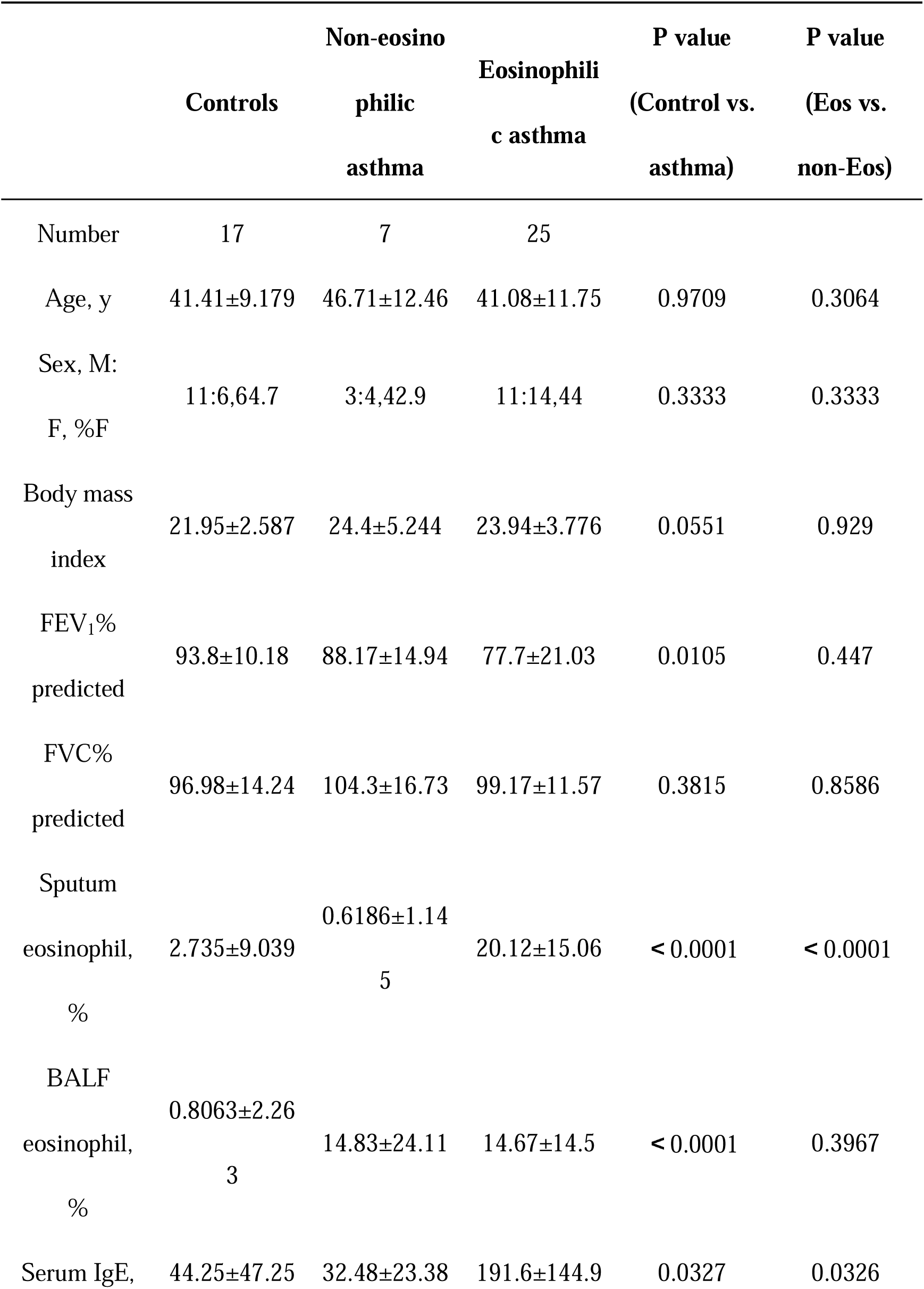
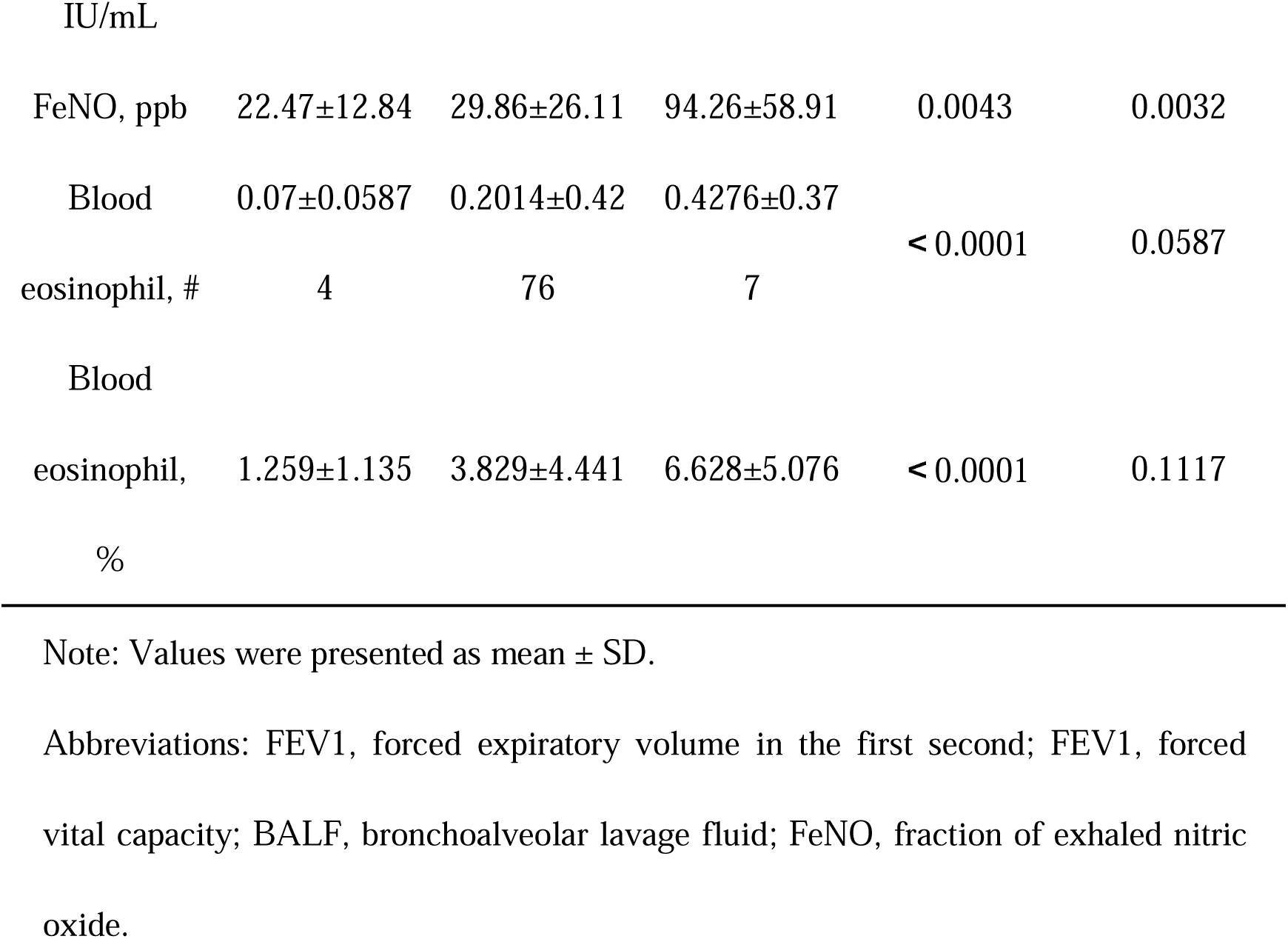
Subject characteristics

**Figure 1.**
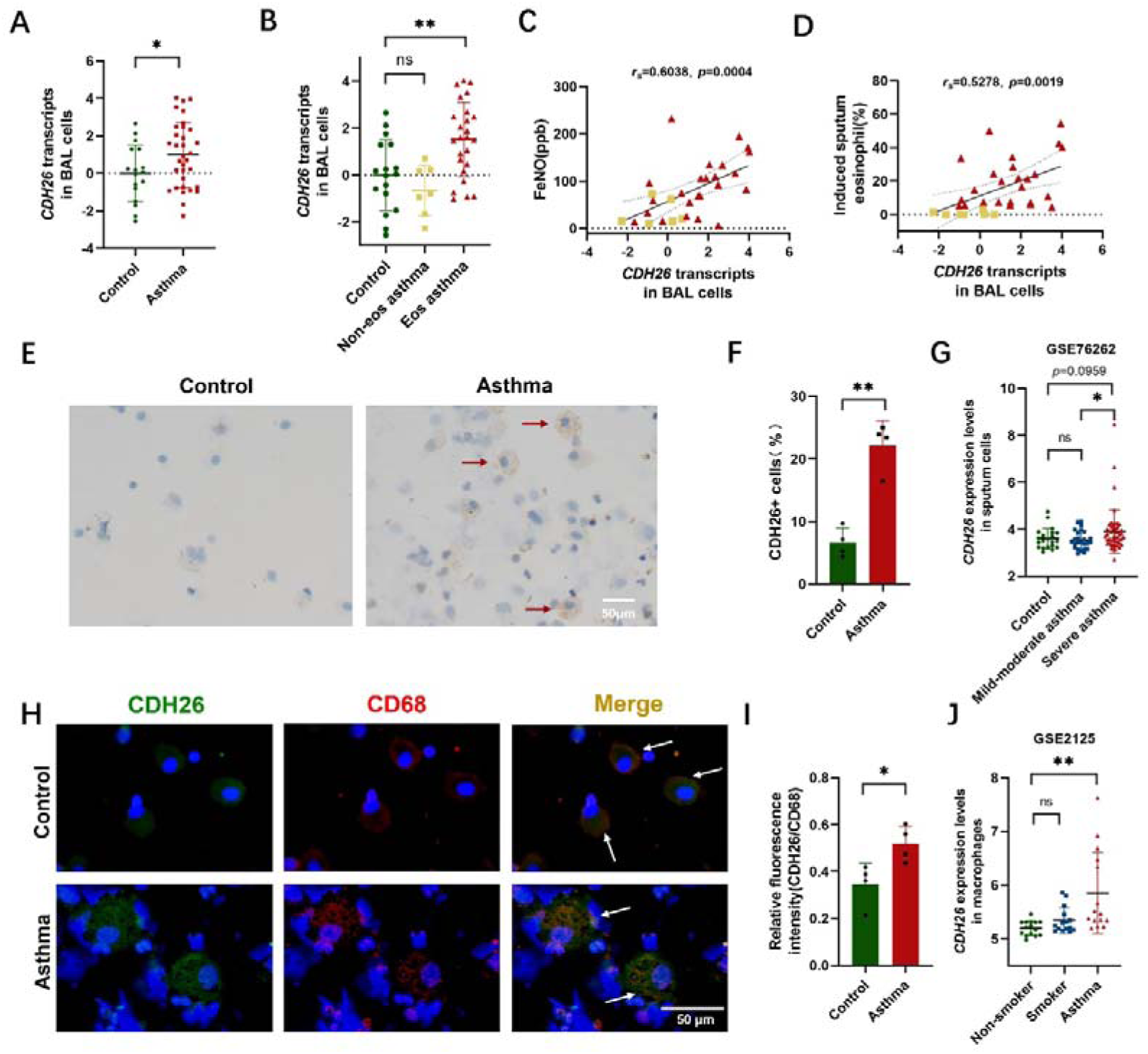
CDH26 expression is upregulated in lung macrophages from eosinophilic asthma patients. A-B) *CDH26* transcript levels in BAL cells from eosinophilic asthmatics (n=25), non-eosinophilic asthmatics (n=7) and control subjects (n=17) were measured by quantitative PCR. The transcript levels were expressed as log2 transformed and relative to the mean value for control subjects. C-D) Spearman’s correlation assays between *CDH26* transcripts in BAL cells and FeNO, induced sputum eosinophil (%) of asthma patients. E) Representative images of CDH26 immumohistochemical staining in BAL cells from asthmatics (n=4) and control subjects (n=4). F) The proportion of CDH26^+^ cells in BAL cells from asthma patients (n=4) and control subjects (n=4). G) The expression levels of *CDH26* in a microarray data (GSE76262) using sputum cells from subjects with mild-moderate (n=25) and severe asthma (n=49) were analyzed. H-I) Representative images for immunofluorescence staining of CDH26 (green) and CD68 (red) in BAL cells from asthma patients (n=4) and control subjects (n=4). Arrows indicate enlarged, foamy, and granular macrophages. Nuclei was stained with DAPI (blue). Quantification for relative immunofluorescent intensity of CDH26 compared to CD68 was performed using ImageJ. J) The expression of CDH26 in isolated alveolar macrophages from asthma patients and control subjects (GSE2125). Data are mean ± SD. \**p*<0.05; \*\**p*<0.01. Abbreviations: CDH26, cadherin-26; FeNO, fraction of exhaled nitric oxide.

We next examined CDH26 expression in BAL cells using immunohistochemistry and found a significant increase in the number of CDH26-expressing cells in BAL cells from asthma patients compared to control subjects (Figures 1E-F). Immunofluorescence staining of BAL cells with CDH26 and CD68, a marker for macrophages, revealed that CDH26 was localized to enlarged foamy CD68-expressing cells, and the fluorescent intensity of CDH26 relative to CD68 was markedly increased in asthma patients (Figures 1H-I). Of note, the CDH26^+^CD68^+^ cells from asthma patients exhibited an enlarged, foamy, and granular morphology, which was previously reported to be associated with alternative activation of macrophages (27). This suggests that CDH26 is expressed in activated macrophages and its expression is upregulated in asthma. We next analyzed a dataset (GSE76262) using sputum cells from mild-moderate and severe asthma, and found that *CDH26* expression levels were higher in severe asthma compared to mild-moderate asthma (Figure 1G) (28). We further analyzed a dataset (GSE2125) using alveolar macrophages isolated from asthma patients and control subjects, and *CDH26* expression levels were significantly higher in macrophages from asthma patients compared to non-smoker controls (Figure 1J) (29). Together, these data suggest that CDH26 expression is upregulated in BAL macrophages of eosinophilic asthma.

### 2. Macrophage-specific *Cdh26* deficiency alleviates airway eosinophilia and mucus overproduction in a mouse model of allergic airway inflammation

We next examined the expression of Cdh26 in BAL cells from wild type (WT) mice and observed enlarged, foamy Cdh26^+^Cd68^+^ cells in in ovalbumin (OVA)-sensitized and challenged WT mice (Figure 2A). The fluorescent intensity of Cdh26 relative to Cd68 was increased in OVA challenged WT mice (Figure 2B). Global *Cdh26* deficiency significantly inhibited airway eosinophilia and the expression of markers for macrophage alternative activation in the mouse model (Figures S2A-C, S3A-I). We further utilized macrophage-specific *Cdh26*-deficient mice generated by crossing *Lyz2Cre* mice and *Cdh26^fl/fl^* mice (Figures S4A-C, 2C). H&E staining and morphological inflammatory scoring revealed that OVA-induced peribronchial inflammation was alleviated in *Cdh26^fl/fl^Lyz2Cre* mice compared to *Cdh26^fl/fl^* mice (Figures 2D-E). OVA challenge increased the number of PAS-staining-positive mucous cells and BAL eosinophils in *Cdh26^fl/fl^* mice, but these increases were significantly reduced in *Cdh26^fl/fl^Lyz2Cre* mice (Figures 2D, 2F, 2G). In addition, Masson trichrome staining revealed that collagen area was decreased in *Cdh26^fl/fl^Lyz2Cre* mice (Figures S5A-B). Our findings suggest that macrophage *Cdh26* contributes to eosinophilic airway inflammation and mucus overproduction in the mouse model of allergic airway inflammation.

**Figure 2.**
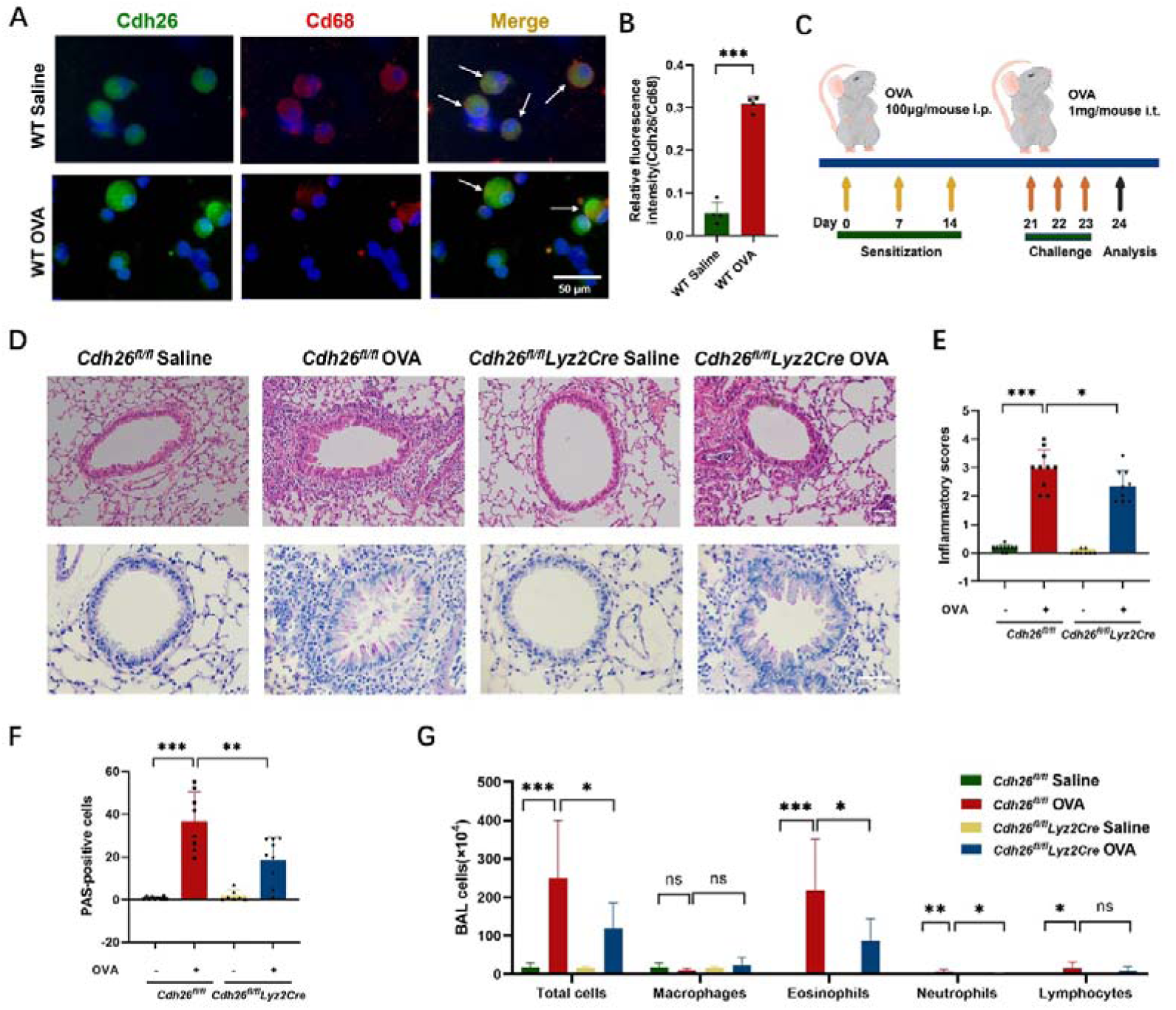
Macrophage-specific *Cdh26* deficiency alleviates airway eosinophilia and mucus overproduction in a mouse model of allergic airway inflammation. A-B) Representative images for co-immunofluorescence staining of Cdh26 (green) and Cd68 (red) in BAL cells from mouse (scale bar = 50 μm). Quantification of relative immunofluorescence intensity of Cdh26 to Cd68 (n=4). C) Mouse experimental schedule. D) Representative images of H&E and PAS staining in mouse lung sections (scale bar = 100 μm). E) The inflammatory score was graded in four random fields for each lung section at 200× magnification. F) The numbers of PAS-staining-positive cells were counted in four random fields for each lung section at 200× magnification. G) Cell counts for macrophages, eosinophils, lymphocytes and neutrophils in BALF. Data are mean ± SD. \**p*<0.05; \*\**p*<0.01; \*\*\**p*<0.001. Abbreviations: Cdh26, cadherin-26; Cd68, Cd68 molecule; WT, wild type; OVA, ovalbumin; H&E staining, hematoxylin-eosin staining; PAS staining, periodic acid-Schiff; BAL, bronchoalveolar lavage.

### 3. Cdh26 contributes to macrophage alternative activation

We further evaluated the effect of macrophage-specific *Cdh26* deficiency on macrophage alternative activation. Immunofluorescence staining for Cd68 and Cd206, a marker for M2 macrophage, was performed using mouse BAL cells. The number of Cd68^+^Cd206^+^ BAL cells was increased in OVA-challenged *Cdh26^fl/fl^* mice while this increase was suppressed in *Cdh26^fl/fl^Lyz2Cre* mice (Figures 3A-B). We also used flow cytometry to evaluate macrophage alternative activation in lung tissue. Defining Cd45^+^Cd11b^+^Cd206^+^ cells as M2 macrophages (Figure S4D), the number of these cells were decreased in lung tissue of OVA-challenged *Cdh26^fl/fl^Lyz2Cre* mice compared to *Cdh26^fl/fl^*mice (Figures 3C-D). Consistently, western blotting showed reduced expression of M2 markers including Arg1 and Cd206 in lung tissue of OVA-challenged *Cdh26^fl/fl^Lyz2Cre* mice (Figures 3E-G). The transcript levels of *Cd206* and *Ym1* were also decreased in lung tissue of *Cdh26^fl/fl^Lyz2Cre* mice (Figures 3H-I). M2 macrophages can recruit Th2 cells and promote eosinophilic airway inflammation by secreting CCL17 and CCL22 (30). We found a significant reduction in BALF Ccl17 and Ccl22 protein levels in OVA-challenged *Cdh26^fl/fl^Lyz2Cre* mice (Figures 3J-K).

**Figure 3.**
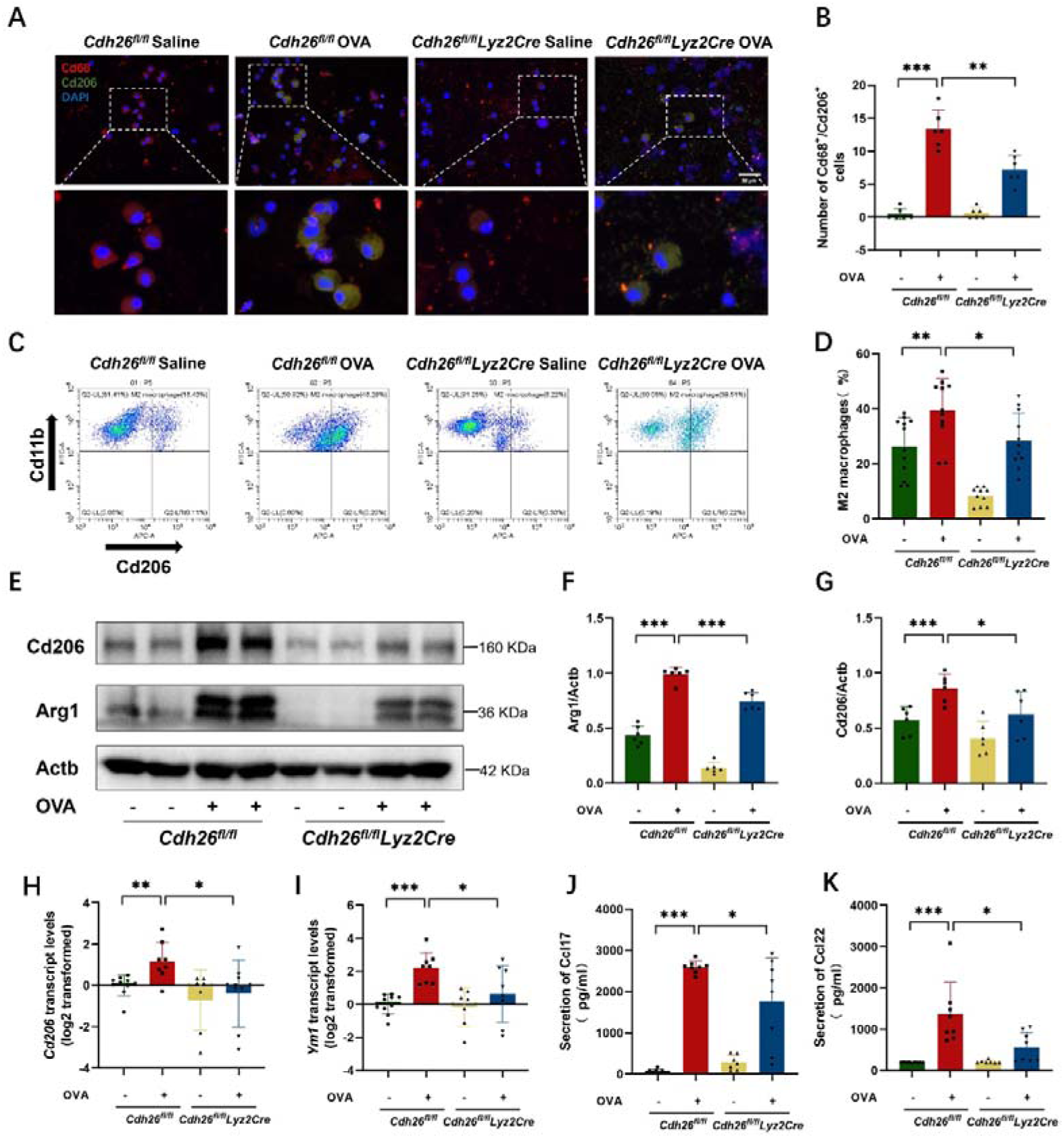
Cdh26 is required for macrophage alternative activation. A) Representative images for immunofluorescence staining of Cd206 (green) and Cd68 (red) in BAL cells from mouse (scale bar = 50 μm). Nuclei was stained with DAPI (blue). B) The numbers of Cd68^+^/Cd206^+^ cells were counted in the cytospin of BAL cells in 5 fields for each mouse and the average number were calculated (n = 6 mice) using ImageJ. C) Flow cytometry for M2 macrophages using single cell suspensions from mouse lung tissue. Cd45^+^Cd11b^+^ cells were identified as macrophages, and Cd45^+^Cd11b^+^Cd206^+^ cells were identified as M2 macrophages. Representative dot plots showing the percentages of M2 macrophages in different groups. D) The percentages of M2 macrophages of total macrophages in different groups. E-G) The protein levels of Arg1, Cd206 in mouse lungs (n=6) were determined by western blotting and representative image was shown. Densitometry assay was performed using ImageJ, and Arg1, Cd206 protein levels were indexed to Actb. H-I) The mRNA levels of *Cd206* and *Ym1* in mouse lungs were determined using quantitative PCR. The transcript levels were expressed as log2 transformed and relative to the mean value of control group. J-K) The protein levels of Ccl17, Ccl22 in the supernatant of BAL fluid were determined using ELISA. n=8-10 mice per group. Data are mean ± SD. \**p*<0.05; \*\**p*<0.01; \*\*\**p*<0.001. Abbreviations: Cd68, Cd68 molecule; Cd206, mannose receptor C-type 1; Arg1, arginase-1; Ym1, also named as Chil3, chitinase-like protein 3; Ccl17, C-C motif chemokine ligand 17; Ccl22, C-C motif chemokine ligand 22; Actb, actin beta.

In vitro, we stimulated primary culture of mouse bone marrow-derived macrophages (BMDMs) and lung macrophages from WT mice with LPS and IL-4, respectively. *Cdh26* transcript levels were increased after IL-4 stimulation, but not altered after LPS stimulation (Figures S6A-F). IL-4 but not IL-13 enhanced *Cdh26* expression in WT BMDMs (Figures S6G-I). Of note, the expression of M2 macrophage markers in IL-4-stimulated BMDMs derived from *Cdh26*^-/-^ mice were significantly suppressed compared to IL-4-stimulated BMDMs from control mice (Figures S7A-I). Similar results were observed in cultured mouse lung macrophages (Figures S8A-E). Our in vivo and in vitro data suggest that *Cdh26* contributes to macrophage alternative activation.

### 4. *Cdh26* deficiency suppresses IL-4Rα expression and IL-4R-STAT6 signaling in macrophages

Type I IL-4R consisting of IL-4Rα and IL-2Rγ (γc) subunits plays a central role in macrophage alternative activation (15). We examined the expression of CDH26 and IL-4Rα in BAL cells using immunofluorescence staining, and found that both CDH26 and IL-4Rα were localized in some CD206^+^ macrophages (Figure 4A), and the number of CDH26^+^IL-4Rα^+^CD206^+^ cells were markedly increased in asthma patients compared to control subjects (Figure 4B). This suggests that CDH26 and IL-4Rα are co-expressed in alternatively activated BAL macrophages in human asthma. Moreover, the transcription levels of *IL4RA* were increased in sputum cells of our mild-moderate asthma patients compared to control subjects (Figure 4C). We analyzed the dataset GSE2125 using human alveolar macrophages and found that *IL4RA* expression levels were higher in asthma compared to control subjects (Figure 4D). This suggests that IL-4Rα is upregulated in alveolar macrophages in asthma. Of note, analysis of the dataset GES76262 revealed that *IL4RA* expression levels were higher in sputum cells of severe asthma compared to controls (Figure 4E).

**Figure 4.**
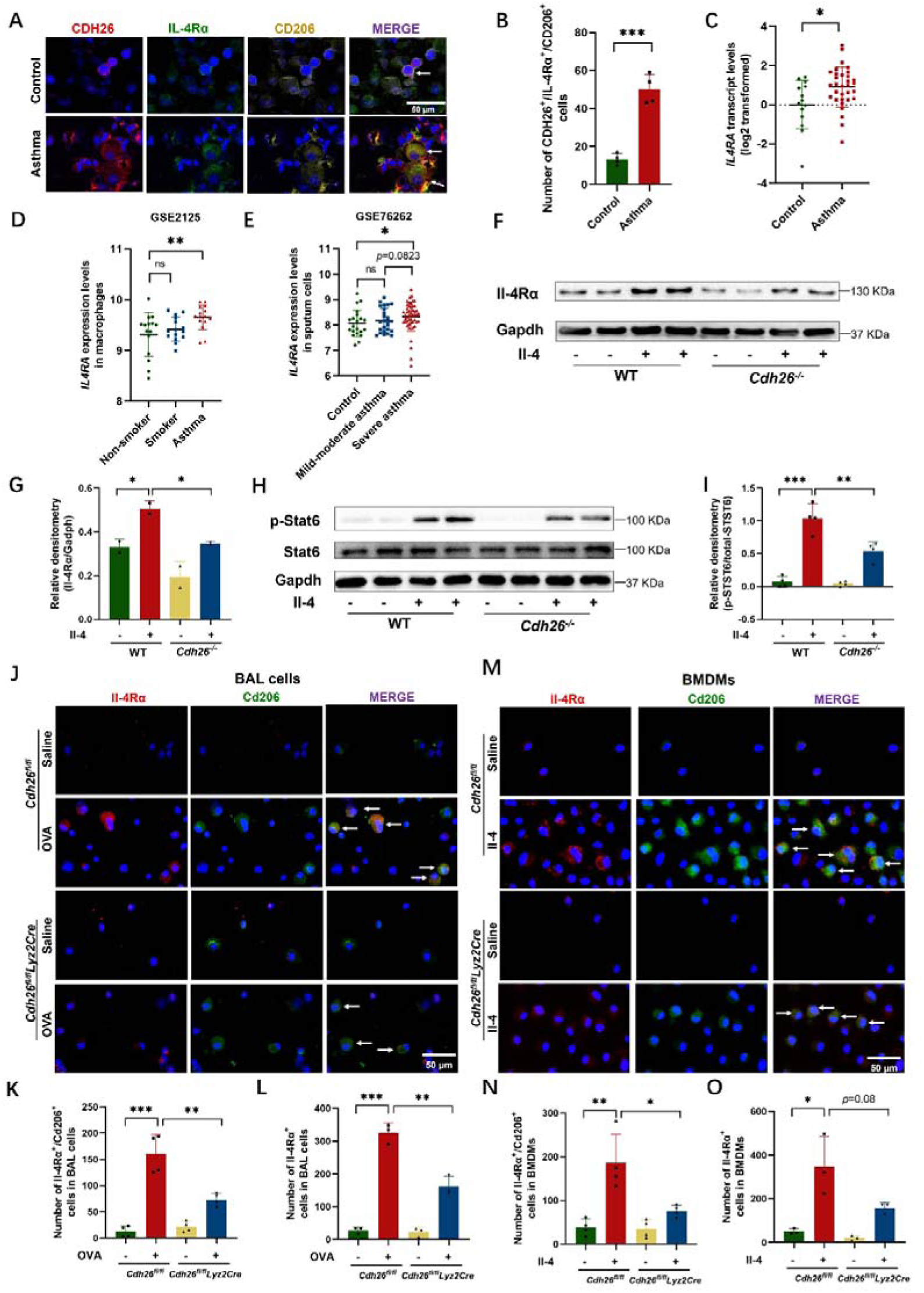
*Cdh26* deficiency suppresses IL-4R expression and IL-4R-STAT6 signaling in macrophages. A) Representative images of immunostaining of CDH26 (red), IL-4Rα (green) and CD206 (yellow) in BAL cells from asthma patients (n=4) and control subjects (n=4). Nuclei was stained with DAPI (blue). B) The numbers of CDH26^+^/IL-4Rα ^+^/CD206^+^ cells were counted in the cytospin of BAL cells in 5 fields for each subject and the average number were calculated using ImageJ (n = 4 per group). C) *IL4RA* transcript levels in induced sputum cells from asthma patients (n=32) and control subjects (n=14) were determined by quantitative PCR. The transcript levels were expressed as log2 transformed and relative to the mean value for control subjects. D) The expression of *IL4RA* in isolated alveolar macrophages from asthma patients and control subjects (GSE2125). E) The expression levels of *IL4RA* in dataset GSE76262 using sputum cells from subjects with mild-moderate (n=25) and severe asthma (n=49) were analyzed. F-G) The protein levels of Il-4Rα in WT or *Cdh26*^-/-^ BMDMs exposed to Il-4 for 48h were determined by western blotting. Densitometry assay was performed using ImageJ, and Il-4Rα protein levels were indexed to Gapdh. H-I) The protein levels of p-STAT6 in WT or *Cdh26*^-/-^ BMDMs stimulated with or without Il-4 for 6h were determined by western blotting. Densitometry assay was performed using ImageJ, and p-STAT6 protein levels were indexed to total STAT6. J) Representative images for immunofluorescence staining of Il-4Rα (green) and Cd206 (red) in BAL cells from *Cdh26^fl/fl^Lyz2Cre* and *Cdh26^fl/fl^* mice challenged with saline or OVA. Nuclei was stained with DAPI (blue). K-L) The numbers of Il-4Rα^+^Cd206^+^ cells (K), Il-4Rα^+^ (L) were counted in the cytospin of BAL cells in 5 fields for each mouse and the average number were calculated for *Cdh26^fl/fl^Lyz2Cre* and *Cdh26^fl/fl^* mice challenged with saline or OVA using ImageJ (n = 4 mice per group). M) Representative images for immunofluorescence staining of Il-4Rα (green) and Cd206 (red) in saline- or Il-4-stimulated BMDMs derived from *Cdh26^fl/fl^Lyz2Cre* and *Cdh26^fl/fl^* mice. Nuclei was stained with DAPI (blue). N-O) The numbers of Il-4Rα^+^Cd206^+^(N), Il-4Rα^+^(O) cells were counted in saline- or Il-4-stimulated BMDMs derived from *Cdh26^fl/fl^Lyz2Cre* and *Cdh26^fl/fl^* mice using ImageJ (n=4 wells per group). Data are mean ± SD. \**p*<0.05; \*\**p*<0.01; \*\*\**p*<0.001. Abbreviations: CDH26, cadherin-26; CD206, mannose receptor C-Type 1; IL-4Rα, interleukin-4 receptor subunit alpha; BAL, bronchoalveolar lavage; BMDM, bone marrow derived macrophage; STAT6, signal transducer and activator of transcription 6; Gapdh, glyceraldehyde-3-phosphate dehydrogenase.

We next examined Il-4Rα expression in cultured BMDMs from WT and *Cdh26*^-/-^ mice using western blotting. *Cdh26* deficiency suppressed recombinant mouse Il-4-induced Il-4Rα protein expression in BMDMs (Figures 4F-G). We further assessed the phosphorylation of STAT6 using western blotting and observed that *Cdh26* deficiency significantly inhibited Il-4-induced STAT6 phosphorylation in BMDMs (Figures 4H-4I). To further investigate the impact of *Cdh26* deficiency on Il-4Rα expression in macrophages, we performed co-immunofluorescence staining of Il-4Rα and Cd206 in mouse BAL cells. OVA challenge increased the number of Il-4Rα^+^ and Il-4Rα^+^Cd206^+^ cells in BAL cells from *Cdh26^fl/fl^* mice, while the number of these cells were decreased in OVA-challenged *Cdh26^fl/fl^Lyz2Cre* mice (Figures 4J-L). Similar results were observed in saline or Il-4-stimulated BMDMs from *Cdh26^fl/fl^* and *Cdh26^fl/fl^Lyz2Cre* mice (Figures 4M-O). Our data suggest that *Cdh26* deficiency suppresses IL-4Rα expression and the IL-4R-STAT6 signaling in macrophages.

### 5. CDH26 enhances IL-4Rα expression via inhibiting its ubiquitination and degradation

To investigate the mechanism by which CDH26 regulates IL-4Rα expression, we used cycloheximide, an inhibitor of protein synthesis, to treat A549 cells which constitutively express IL-4Rα, and assessed the impact of CDH26 knockdown or overexpression on the degradation rate of IL-4Rα and IL-2Rγ. CDH26 knockdown significantly accelerated the degradation rate of IL-4Rα whereas did not alter that of IL-2Rγ (Figures 5A-C). Conversely, CDH26 overexpression significantly suppressed the degradation rate of IL-4Rα whereas did not alter that of IL-2Rγ (Figures 5D-F). This indicates that CDH26 suppresses IL-4Rα degradation.

**Figure 5.**
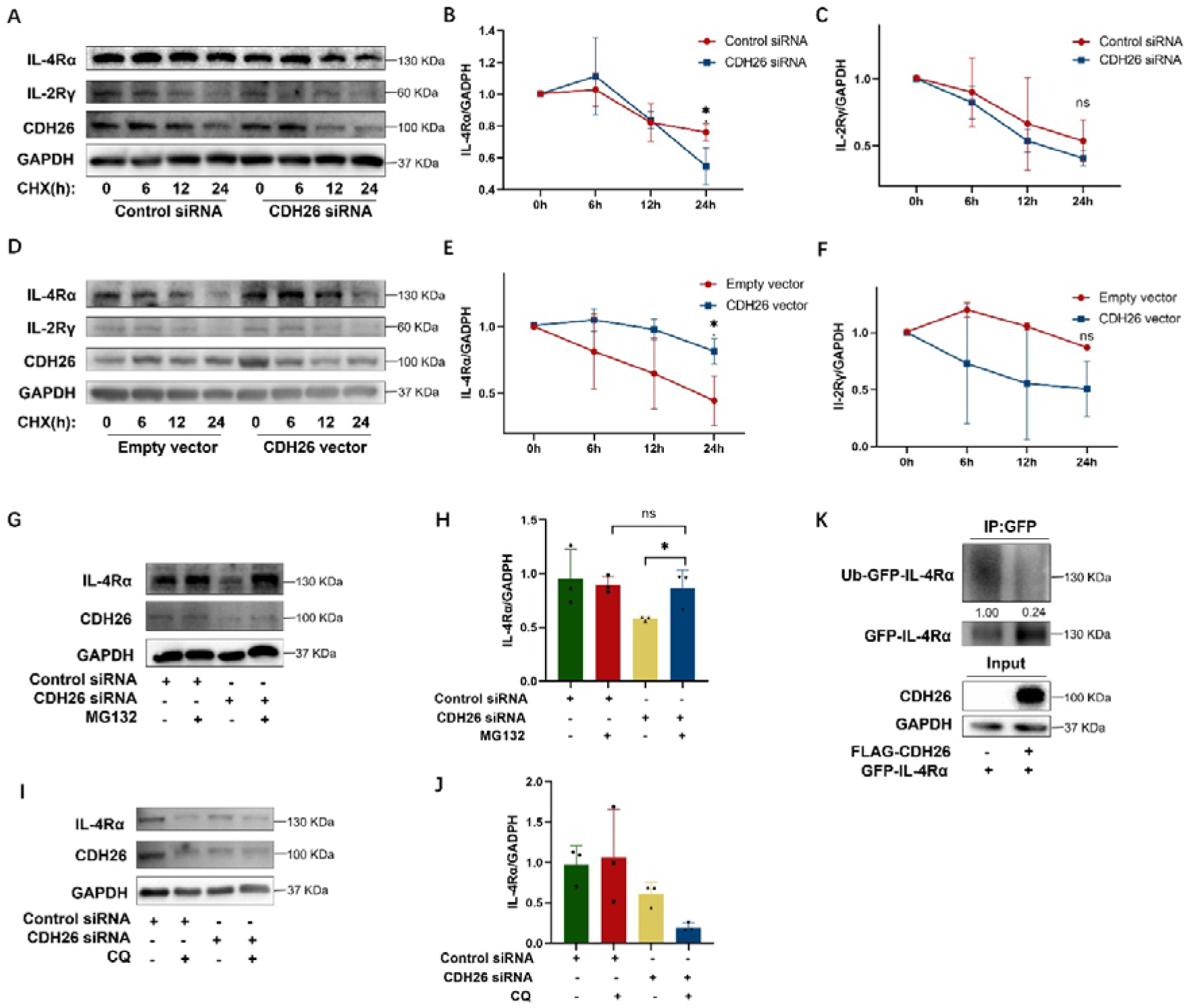
CDH26 enhances IL-4R_α_ expression via inhibiting its ubiquitination and degradation. A-C) IL-4Rα and IL-2Rγ protein levels were measured in A549 cells transfected with control or CDH26 siRNA and treated with CHX for 0, 6, 12, 24h using western blotting. Densitometry assay was performed using ImageJ, and IL-4Rα and IL-2Rγ protein levels were indexed to GAPDH. D-F) IL-4Rα and IL-2Rγ protein levels were measured in A549 cells transfected with empty or CDH26 overexpressing vector and treated with CHX for 0, 6, 12, 24h using western blotting. Densitometry assay was performed using ImageJ, and IL-4Rα and IL-2Rγ protein levels were indexed to GAPDH. The experiment was repeated 3 times independently. G-H) IL-4Rα protein level was determined in A549 cells transfected with control or CDH26 siRNA and treated with MG132 using western blotting. Densitometry assay was performed using ImageJ, and IL-4Rα protein levels were indexed to GAPDH. I-J) IL-4Rα protein level was determined in A549 cells transfected with control or CDH26 siRNA and treated with CQ using western blotting. Densitometry assay was performed using ImageJ, and IL-4Rα protein level was indexed to GAPDH. K) HEK-293T cells were transfected with plasmids expressing IL-4Rα^GFP^ and CDH26^FLAG^ as indicated, and cell lysates were immunoprecipitated with anti-GFP antibody followed by western blotting. The ubiquitination level of IL-4Rα was assessed using ubiquitin antibodies. All experiments were repeated for at least three times independently. Data are mean ± SD. \**p*<0.05. Abbreviations: CDH26, cadherin-26; IL-4Rα, interleukin-4 receptor subunit alpha; IL-2Rγ, interleukin-2 receptor subunit gamma; GAPDH, glyceraldehyde-3-phosphate dehydrogenase; CHX, cycloheximide; MG132, Z-Leu-Leu-Leu-al; CQ, <u>chloroquine</u>.

Protein degradation pathways include the ubiquitin-proteasome pathway and the lysosome pathway. To investigate the specific mechanism by which CDH26 suppresses IL-4Rα degradation, we used the proteasome pathway inhibitor MG132 and the lysosome pathway inhibitor chloroquine to treat A549 cells transfected with CDH26 siRNA. We found that CDH26 knockdown decreased IL-4Rα protein level while MG132 restored the protein level of IL-4Rα. However, chloroquine did not affect IL-4Rα protein expression (Figures 5G-J). These results suggest that CDH26 may inhibit IL-4Rα degradation through the ubiquitin-proteasome pathway. In support of this, we found that CDH26 overexpression significantly inhibited the ubiquitination level of IL-4Rα (Figures 5K). Together, our findings suggest that CDH26 enhances IL-4Rα expression via inhibiting its ubiquitination and degradation.

### 6. CDH26 interacts with STUB1 to restrain the binding of STUB1 with IL-4Rα

To further explore the mechanism by which CDH26 inhibits IL-4Rα ubiquitination-proteasomal degradation, we transfected HEK-293T cells with CDH26-Flag overexpression plasmid. Co-immunoprecipitation experiment using Flag antibody and mass spectrometry analysis were performed to identify the proteins interacting with CDH26 (Figures 6A, S9A-B). A total of 423 proteins interacting with CDH26 were identified and they were intersected with the molecules involved in “Ubiquitin mediated proteolysis” pathway in KEGG database, resulting in the identification of four molecules: UBE3C, STUB1, CDC20, and UBE2G2 (Figures 6B-C). A previous study demonstrated that STUB1 acts as an E3 ubiquitin ligase responsible for IL-4Rα ubiquitination and subsequent degradation (17). *STUB1* transcripts were slightly decreased in BAL cells but not altered in airway brushings from asthma patients compared to control subjects (Figures 6D-E). There was no significant difference in Stub1 transcript and protein levels in lung tissues between OVA-challenged *Cdh26^fl/fl^Lyz2Cre* and control mice (Figures 6F-H).

**Figure 6.**
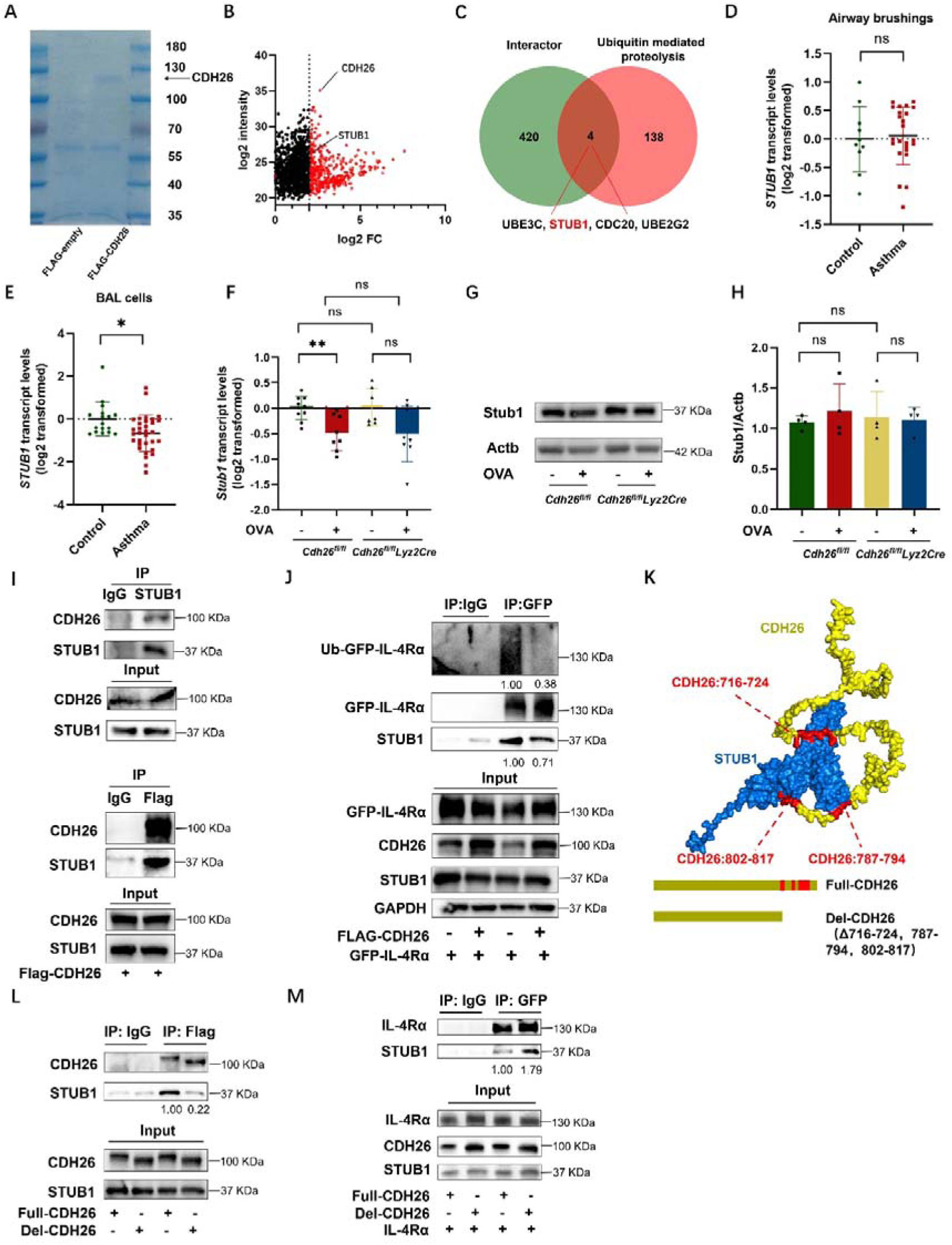
CDH26 interacts with STUB1 to restrain the binding of STUB1 with IL-4R_α_. A) HEK-293T cells were transfected with plasmids expressing CDH26^FLAG^, and cell lysates were immunoprecipitated with anti-FLAG antibody followed by mass spectrum. B) Volcanic map of the proteins interacting with CDH26 detected by mass spectrum. C) Veen diagram shown the protein molecules identified by intersecting the proteins interacting with CDH26 and the proteins involved in ubiquitin-mediated proteolysis. D) *STUB1* transcript levels in airway brushings from asthma patients (n=24) and control subjects (n=10) were determined using quantitative PCR. E) *STUB1* transcript levels in BAL cells from asthma patients (n=32) and control subjects (n=17) were determined using quantitative PCR. F) The transcript levels of *Stub1* in lungs of *Cdh26^fl/fl^Lyz2Cre* or *Cdh26^fl/fl^* mice challenged with saline or OVA were determined using quantitative PCR (n = 8-10 mice per group). The transcript levels were expressed as log2 transformed and relative to the mean value of control group. G-H) The protein levels of Stub1 in lungs of *Cdh26^fl/fl^Lyz2Cre* or *Cdh26^fl/fl^* mice challenged with saline or OVA were determined using western blotting (n = 3 per group). Densitometry assay was performed using ImageJ, and Stub1 protein levels were indexed to Actb. I) Binding of CDH26 with STUB1 was examined using immunoprecipitation assay. J) HEK-293T cells were transfected with plasmids expressing IL-4Rα^GFP^ and CDH26^FLAG^ as indicated, and cell lysates were immunoprecipitated with anti-GFP or IgG antibody followed by western blotting. The ubiquitination level of IL-4Rα was assessed using ubiquitin antibodies. K) The binding interface between CDH26 (716-724, 787-794, and 802-817) and STUB1 was predicted based on the molecular docking model using zdock. L) HEK-293T cells were transfected with plasmids expressing full-CDH26 (1–832) and del-CDH26^FLAG^ (deleting amino acids 716-724, 787-794, and 802-817), and cell lysates were immunoprecipitated with anti-FLAG or IgG antibody followed by western blotting. M) HEK-293T cells were transfected with plasmids expressing IL-4Rα^GFP^, full-CDH26 and del-CDH26^FLAG^, and cell lysates were immunoprecipitated with anti-GFP or IgG antibody followed by western blotting. Data are mean ± SD. \**p*<0.05. Abbreviations: CDH26, cadherin-26; STUB1, STIP1 homology and U-box containing protein 1; ACTB, actin beta; GAPDH, glyceraldehyde-3-phosphate dehydrogenase; IL-4Rα, interleukin-4 receptor subunit alpha.

We further confirmed the interaction between CDH26 and STUB1 using protein immunoprecipitation experiments. Coimmunoprecipitation assay using STUB1 antibody to capture protein complexes in HEK-293T lysates showed that CDH26 coprecipitated with STUB1 (Figure 6I). In reciprocal experiments, overexpression of Flag-tagged CDH26 in HEK-293T cells resulted in the coprecipitation of STUB1 with the antibody against Flag (Figure 6I). To explore the impact of the CDH26-STUB1 interaction on the ubiquitination of IL-4Rα, we transfected HEK-293T cells with a plasmid overexpressing CDH26 and observed a significant reduction of STUB1 levels binding to IL-4Rα, as well as a decrease in the ubiquitination level of IL-4Rα (Figure 6J). This suggests that CDH26 directly interacts with STUB1, inhibiting the binding of STUB1 with IL-4Rα and its subsequent ubiquitination. Next, we determined the potential binding sites of CDH26 required for its interaction with STUB1 as described in Methods. We designed CDH26 mutants by deleting the amino acids 716-724, 787-794, and 802-817 which putatively bind to STUB1. Protein immunoprecipitation experiments revealed that deletion of these segments significantly weakened the binding of CDH26 to STUB1, while the binding of STUB1 to IL-4Rα was significantly enhanced (Figures 6L-M). Our data suggest that CDH26 interacts with STUB1 to restrain the binding of STUB1 with IL-4Rα.

### 7. *Cdh26* siRNA encapsulated lipid nanoparticles alleviate airway eosinophilia and mucus overproduction in the mouse model

Finally, we examined the possibility of Cdh26 as a therapeutic target for asthma. We screened for efficient *Cdh26* siRNAs and encapsulated *Cdh26* siRNA-2 in lipid nanoparticles (LNPs) (Figure 7A). The encapsulation efficiency of *Cdh26* siRNA@LNPs was 95.4% and the particle size was 86.2nm (Table S2). After administration of DiOC_18_ (DiR)-labeled LNPs intratracheally, we found that DiR-labeled LNPs were present in Cd68^+^ BAL cells (Figures 7B). Using in vivo imaging system, we found that the fluorescent signal lasted for at least 3 days in mouse lungs (Figure 7C). In control and *Cdh26* siRNA@LNPs treated mouse models (Figure 7D), H&E staining of liver, spleen, and kidney showed no significant damage (Figure S10). Immunofluorescence staining of Cdh26 and Cd68 in BAL cells revealed decreased Cdh26 expression in Cd68^+^ cells after *Cdh26* siRNA@LNPs treatment (Figure 7F). *Cdh26* siRNA@LNPs treatment significantly decreased the number of BAL eosinophils, alleviated peribronchial inflammation, and suppressed mucous cell metaplasia (Figures 7E-H). Moreover, *Cdh26* siRNA@LNPs treatment inhibited the expression of markers for macrophage alternative activation including Arg1, Ym1, Ccl17, and Ccl24 (Figures 7I-M). Our data suggest that *Cdh26* siRNA encapsulated lipid nanoparticles alleviate airway eosinophilia and mucus overproduction in the mouse model.

**Figure 7.**
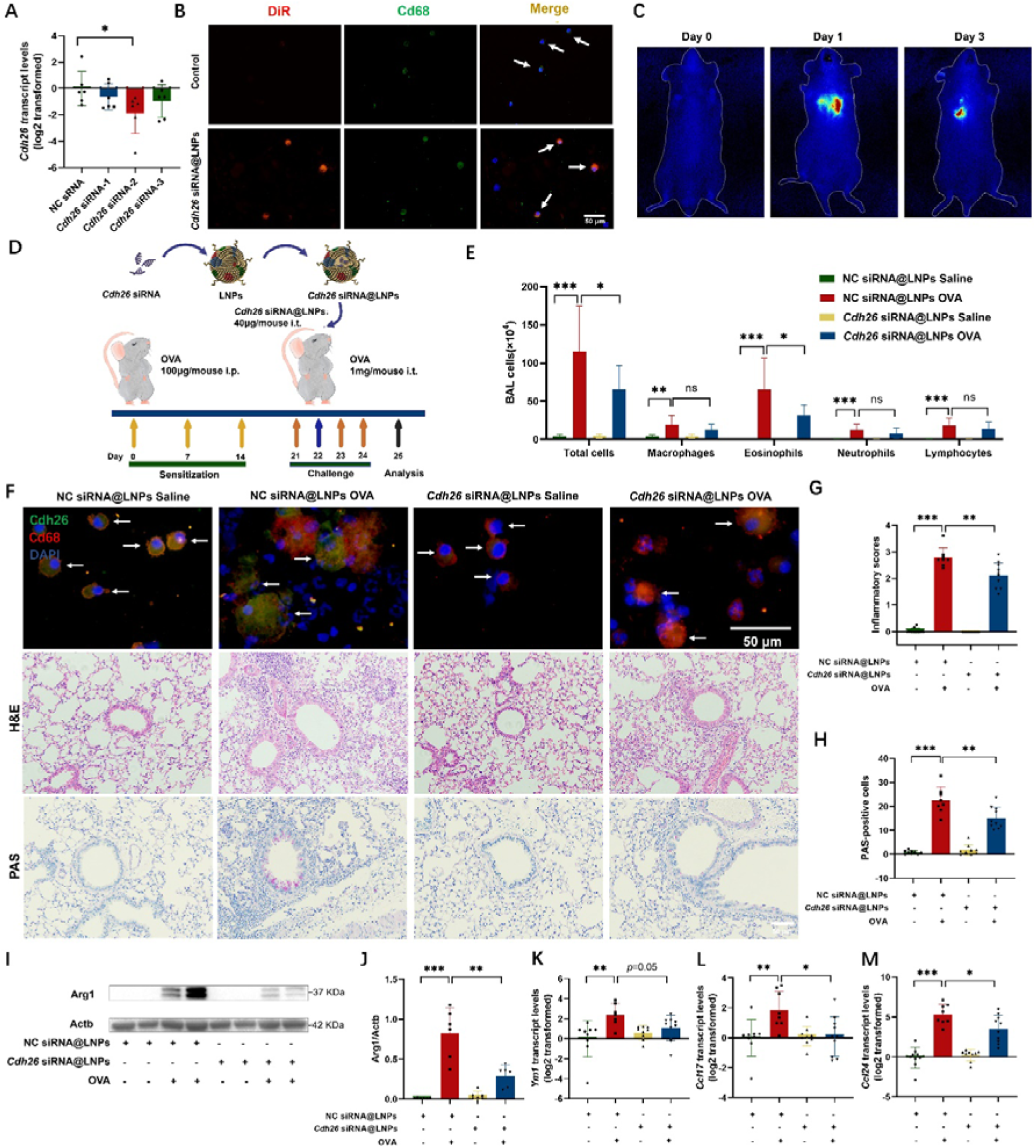
*Cdh26* siRNA encapsulated lipid nanoparticles effectively alleviates airway eosinophilia and mucus overproduction in the mouse model. A) *Cdh26* transcripts in 3T3 cells transfected with three Cdh26 siRNA were measured by quantitative PCR. The transcript levels were expressed as log2 transformed and relative to the mean value for control group. B) Representative images for immunofluorescence staining of DiR (red) and Cd68 (green) in mouse BAL cells. C) Representative images obtained by in vivo image system for mice at days 0, 1, and 3 after administration with DiR-labeled LNPs. D) Mouse experimental schedule. E) Cell counts for macrophages, eosinophils, lymphocytes and neutrophils in mouse BALF. F) Representative images of immunofluorescence staining of Cdh26 (green) and Cd68 (green) in BAL cells, H&E staining and PAS staining in mouse lung sections. G) The inflammatory score was graded in four random fields for each lung section at 200× magnification. H) The numbers of PAS-staining-positive cells were counted in four random fields for each lung section at 200× magnification. I-J) The protein levels of Arg1 in mouse lungs were determined by western blotting, and Arg protein levels were indexed to Actb. K-M) The transctipt levels of *Ym1*, *Ccl17* and *Ccl24* in mouse lungs were determined using quantitative PCR. The transcript levels were expressed as log2 transformed and relative to the mean value for control group. n=8-10 mice per group. Data are mean ± SD. \**p*<0.05; \*\**p*<0.01; \*\*\**p*<0.001. Abbreviations: CDH26, cadherin-26; BALF, bronchoalveolar lavage fluid; LNPs, lipid nanoparticles; DiR, DiOC_18_; Cd68, Cd68 molecule; Ccsp, club cells 10 kDa secretory protein; Arg1, arginase-1; Ym1, also named as Chil3, chitinase-like protein 3; Ccl24, C-C motif chemokine ligand 24; Ccl17, C-C motif chemokine ligand 17.

**Figure 8.**
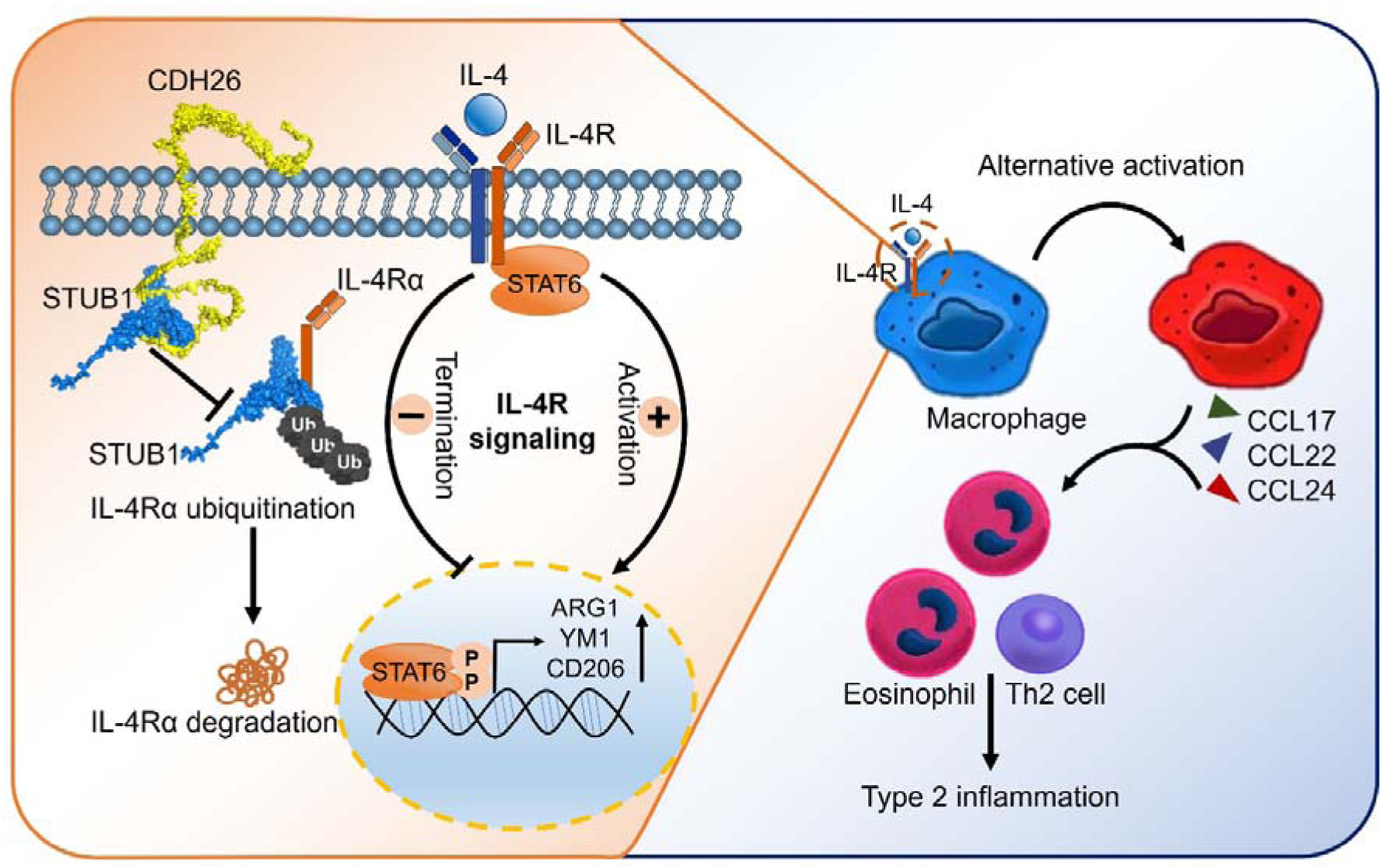
Schematic diagram for the mechanism by which CDH26 amplifies IL-4R signaling. IL-4R signaling is activated by type 2 cytokines including IL-4 and IL-13. STUB1 interacts with IL-4R subunit IL-4Rα and results in its ubiquitination and subsequent degradation, leading to the termination of IL-4R signaling. CDH26, through interacting with STUB1 and trapping it, inhibits IL-4Rα ubiquitination-proteasomal degradation, thus hampering the termination of IL-4R-STAT6 signaling, and resulting in macrophage alternative activation, and type 2 inflammation in asthma.

## Discussion

IL-4R signaling drives mucous cell metaplasia, macrophage alternative activation, Th2 cell differentiation, and type 2 inflammation in allergic diseases (31–36). However, the mechanism underlying the regulation of IL-4R expression remains unclear. Our study uncovers a novel mechanism that CDH26 directly interacts with STUB1 and suppresses the STUB1-mediated ubiquitination-proteasomal degradation of IL-4Rα, thus hampering the termination of IL-4R signaling and resulting in sustained activation of IL-4R signaling in asthma.

As a member of the cadherin family, CDH26 is highly expressed in allergic airway and gastrointestinal epithelium (24, 25). For the first time, we showed that CDH26 was upregulated in activated BAL macrophages from eosinophilic asthma patients. Moreover, CDH26 expression in sputum cells is higher in severe asthma compared to mild-moderate asthma, implicating a role of CDH26 in severe asthma. Several studies suggest a link between increased number of M2 macrophages and asthma severity (37–39). M2 macrophages can release chemokines such as CCL17 and CCL22 to recruit Th2 cells in asthma (40). Our data demonstrated that macrophage-specific *Cdh26* deficiency suppresses airway eosinophilia, macrophage alternative activation and their secretion of CCL17 and CCL22, suggesting that CDH26-mediated macrophage alternative activation contributes to asthma pathogenesis.

IL-4R is a pivotal therapeutic target for asthma patients including those with severe asthma. Our study demonstrated that IL-4Rα expression is upregulated in BAL macrophages from asthma patients, and *Cdh26* deficiency suppresses IL-4Rα expression and downstream STAT6 signaling. The type 2 cytokines IL-4 and IL-13 activate IL-4R signaling in airway epithelial cells, macrophages, and lymphocytes, leading to mucous cell metaplasia, macrophages alternative activation, Th2 cell differentiation, respectively (12). We recently reported that CDH26 promotes mucus cell metaplasia and chemokine secretion by airway epithelial cells via amplifying the type II IL-4R signaling (25). Airway epithelial cells mainly express type C IL-4R consisted of IL-4Rα and IL-13Rα1, while macrophages express type C IL4R composed of IL-4Rα and IL-2Rγ (14). Here, we showed that CDH26 suppresses the degradation of IL-4Rα but not IL-2Rγ. Taken together, enhanced CDH26 inhibits the protein degradation of IL-4Rα, a subunit shared by type I and type II IL-4R, thus promoting macrophage alternative activation and mucous cell metaplasia in asthma.

So far, the mechanisms underlying the regulation of IL-4R expression include endocytosis, ubiquitination-proteasome degradation, and co-receptor system (16–18, 41). Ubiquitin molecules interact with protein substrates through the action of E1 ubiquitin-activating enzymes, E2 ubiquitin-conjugating enzymes, and E3 ubiquitin ligases (42). Wei and colleagues demonstrated the interaction between STUB1, an E3 ubiquitin ligase, and IL-4Rα promotes the ubiquitin-proteasomal degradation of IL-4Rα. *Stub1*-deficient mice exhibit spontaneous airway inflammation and macrophages alternative activation (17). In the present study, we demonstrated that CDH26 directly interacts with STUB1 and suppresses the binding of STUB1 to IL-4Rα, thereby restraining STUB1-mediated IL-4Rα ubiquitination and proteasomal degradation. Our findings provide new insights into the regulation of IL-4R expression.

The highly expressed CDH26 in airway epithelium and macrophages of asthma patients is a promising therapeutic target. Recent studies on lipid nanoparticles have shown that modified lipid nanoparticles can effectively penetrate the mucus-cilia barrier maintaining high stability in nebulized form (43). Our lipid nanoparticles encapsulating *Cdh26* siRNA effectively suppressed airway eosinophilia, mucous cell metaplasia and macrophage alternative activation in a mouse model. Lipid nanoparticles encapsulating *CDH26* siRNA may represent a new method for treating asthma.

Our study has several limitations. First, our asthma patients are mild-moderate asthma, the role of CDH26 in severe asthma needs to be further investigated. Second, perfect macrophage-specific gene deletion systems are not currently available, the *Lyz2Cre* driven gene deletion system we used impacts myelomonocytic cells including monocytes, granulocytes, and macrophages (44). Third, A549 or HEK-293T cells were used in the mechanistic experiments because we could not efficiently knock down or overexpress CDH26 in the primary culture of macrophages. Lastly, the efficacy and safety of LNPs encapsulating *Cdh26* siRNA and its aerosolized form in asthma require further study.

In summary, we demonstrated that CDH26 directly interacts with STUB1 and restrains the binding of STUB1 to IL-4Rα, thereby suppressing IL-4Rα ubiquitination-proteasomal degradation, leading to sustained activation of IL-4R signaling and macrophage alternative activation in asthma. CDH26 is a promising therapeutic target for asthma.

## Author contributions

Study concept: G. Zhen; study design: G. Chen and G. Zhen; acquisition of data: G. Chen, S. Chen, C. Huang, W. Gu, H. Jie, L. Zhao, W. Kong, J. Gao, Y. Feng, L. Yi, and G. Zhen; analysis and data interpretation: G. Chen, S. Chen, P. Gao, and G. Zhen; revision of manuscript: G. Chen, P. Gao, and G. Zhen; manuscript writing: G. Chen and G. Zhen.

## Conflict of interest

The authors have no potential conflicts of interest to disclose.

## Support Statement

This work is supported by National Natural Science Foundation of China (grant 82170036), Key Research and Development Program of Hubei Province (grant 2024BCB280), Innovation and Translation Project of Tongji Hospital (grant 2023CXZH006), Natural Science Foundation of Hubei Province (grant 2024AFB622), Research Foundation of Tongji Hospital (grant 2023B06)

## Data Availability

All data produced in the present study are available upon reasonable request to the authors.

## Supplementary Materials and Methods

### Subjects

We recruited 17 control subjects, 25 eosinophilic asthma patients (sputum eosinophils > 3%), and 7 non-eosinophilic asthma patients (sputum eosinophil ≤ 3%) (1). All participants were Chinese and recruited from Tongji Hospital. None of the participants had a history of smoking or received inhaled or oral corticosteroids or leukotriene antagonists. For each participant, we collected demographic information and induced sputum samples. We also measured spirometry, fractional exhaled nitric oxide (FeNO), and serum IgE levels. All asthma patients had a history of episodic wheezing and met the criteria for bronchial hyperresponsiveness, with a methacholine PD20 of less than 2.505 mg or a 12% or greater; increase in forced expiratory volume in the first second (FEV1) after the inhalation of 200 µg of salbutamol. All participants also underwent bronchoalveolar lavage fluid (BALF) collection. The control subjects had no history of chest tightness or wheezing, and lung function stimulation tests were negative. All participants provided written informed consent, and the study was approved by the ethics committee of Tongji Hospital, Huazhong University of Science and Technology.

### Murine model of allergic airway inflammation

*Cdh26^fl/fl^Lyz2Cre* mice on a C57BL/6N background were constructed from the GemPharmatech (Nanjing, China). Exon 4 of *Cdh26* gene was knocked out in *Cdh26^fl/fl^Lyz2Cre* mice. All experimental mice were sensitized three times on days 0, 7, and 14 by intraperitoneal injection of sensitizing agents (OVA + aluminum hydroxide mixture) at a volume of 200 μl (containing 100 μg OVA). From days 21 to 23, each mouse was intranasally stimulated with 50 μl of OVA solution (containing 1 mg OVA) daily (2). Pulmonary resistance in response to various intravenous methacholine concentrations was measured using the forced oscillation technique with the FlexiVent system (SCIREQ) 24 hours after the final challenge. Bronchoalveolar lavage fluid (BALF) was collected and cell counts for macrophages, eosinophils, lymphocytes, and neutrophils were determined as previously described (3). Lung tissues were collected for histological analysis, quantitative PCR, western blotting, flow cytometry, and immunostaining. All experimental procedures were approved by the Animal Care and Use Committee of Tongji Hospital, Huazhong University of Science and Technology.

### Cell culture and treatment

BMDM (bone marrow-derived macrophages) cells and mouse pulmonary macrophages were isolated from both WT (wild-type) and *Cdh26*^-/-^ mice. They were then treated with IL-4 for 48 hours. HEK-293T and A549 cells were cultured in DMEM medium supplemented with 10% FBS (Biological Industries, Israel). These cells were transfected with either scrambled control or CDH26 siRNA (80 nM; GeneCopoeia) or empty or CDH26 cDNA expression vector (500 ng/mL) using Lipofectamine 3000 (Invitrogen, CA).

### Quantitative PCR

Total RNA was isolated from induced sputum cells, BAL cells, mouse lungs, mouse pulmonary macrophages and BMDM cells using TRIzol (Takara, Tokyo, Japan). The isolated RNA was then reverse transcribed into cDNA using the PrimeScript RT reagent kit (Takara, Tokyo, Japan). The primer sequences for gene amplification were obtained from the primer bank and can be found in Table S1. The expression levels of these genes were determined using the 2−ΔΔCT method. The gene expression values were expressed as log2 transformed and relative to the median expression level of the control group.

### Western blotting

The protein sample preparation involved several steps. Firstly, the cells were lysed using ice-cold RIPA lysis buffer (Servicebio, Wuhan, China) supplemented with EDTA-free Protease Inhibitor Cocktail Tablets. The lysates were then subjected to SDS-PAGE, either 8% or non-reducing, for protein separation. The proteins were subsequently transferred onto a PVDF membrane (Roche, Mannheim, Germany) by electrophoresis. Next, the PVDF membranes were blocked using a solution containing Tris-buffered saline with Tween-20 and nonfat milk. The membranes were then incubated overnight at 4 °C with specific antibodies at a dilution of 1:1000 in 5% milk. Antibodies used in western blotting were: anti-CDH26 mAb (Sigma-Aldrich, 1:1000 dilution), mouse anti-Il-4Rα mAb (R&D system, 1:1000 dilution), human anti-IL-4Rα mAb (Abcam, 1:1000 dilution), anti-IL-2Rγ mAb (Abclonal, 1:1000 dilution), anti-total and phosphorylated Stat6 pAb (Cell Signaling, 1:1,000 dilution), anti-STUB1 mAb (Abcam, 1:1000 dilution), mouse anti-Cd206 mAb (R&D system, 1:1000 dilution) and anti-ARG1 mAb (Proteintech, 1:1000 dilution), anti-FLAG mAb (Proteintech, 1:2000 dilution), anti-GFP mAb (Proteintech, 1:2000 dilution), anti-ubiquitin pAb (Proteintech, 1:1000 dilution), anti-anti-GAPDH pAb (Proteintech, 1:4000 dilution) and anti-anti-ACTB pAb (Proteintech, 1:4000 dilution). After the overnight incubation, the membranes were treated with a horseradish peroxidase-conjugated goat anti-rabbit secondary antibody for 1 hour at room temperature. Finally, the protein signals were detected using an ECL kit according to the manufacturer’s instructions.

### Histology staining

The left lungs of mice were fixed by inflation and embedded in paraffin. 5-μm sections were then cut from the embedded tissues. These sections were subjected to H&E staining, PAS staining, and Masson staining. For H&E staining, lung sections were treated with hematoxylin and eosin staining kit (Servicebio, Wuhan, China) to evaluate peri-bronchial inflammation. The severity of inflammation surrounding airways with a luminal diameter of 170 ± 40 μm in H&E-stained lung sections was assessed using the following scoring system: 0 for normal; 1 for few cells; 2 for a single layer of inflammatory cells; 3 for 2-4 layers of inflammatory cells; and 4 for more than 4 layers of inflammatory cells. For PAS staining, lung sections were stained using a periodic acid-Schiff staining kit (Servicebio, Wuhan, China) to detect mucus. The number of PAS-positive cells in airways with a luminal diameter of 170 ± 40 μm was counted in five randomly selected fields per lung section at 200× magnification.

### Immunofluorescence

The immunofluorescence method of frozen slides was as follows: After the slides were restored to room temperature, they were washed 3 times with PBS for 5 min each time. The immunostaining permeabilization buffer was added and incubated at room temperature for 20 min. Add goat serum blocking buffer and block for 30 minutes. Subsequently, the slices were incubated with primary antibodies in a wet box overnight at 4°C. Antibodies used in immunofluorescence were: anti-CDH26 mAb (Sigma-Aldrich, 1:100 dilution), mouse anti-Il-4Rα mAb (R&D system, 1:200 dilution), human anti-IL-4Rα mAb (R&D 1:200 dilution), anti-CD206 mAb (Proteintech, 1:2000 dilution) and anti-CD68 mAb (Proteintech, 1:2000 dilution). Antibody wash buffer was added to remove the primary antibody, and then the Cy3 or FITC conjugated secondary antibodies (donkey-anti mouse or rabbit IgG, Abbkine) were added and incubated at room temperature for 1 h. After PBS cleaning, DAPI staining is used. Washing again, the slices were sealed with anti-fluorescence quenching sealing agent. Then, slices were observed under fluorescence microscope within one week to avoid fluorescence quenching, and images were collected.

### Flow cytometry

Following euthanasia, the lung samples of mice were dissected in HBSS buffer. Single-cell suspensions were prepared by filtering the samples through a 100 μm nylon filter strainer and washing them in HBSS buffer supplemented with 2% FBS, 20 mM HEPES, and 5 mM EDTA. The cells were then resuspended at a concentration of 1*10^6 cells/100 µL. Subsequently, 100 µL of the cell suspension was added to a flow tube, and 2 µL of FcBlock was added to each tube, followed by incubation on ice for 10 min. Three types of tubes were set up: a blank tube, a single staining tube, and a sample tube. In the single staining tube, a single antibody was added. In the sample tube, 0.5 µL of LIVE/DEAD™ Fixable Yellow Dead Cell Stain Kit (Invitrogen, USA), 2 µL of anti-Cd45 (bv510), and 1.25 µL of anti-Cd11b (FITC) antibodies (Biolegend, USA) were added, followed by incubation on ice for 15-20 min while protected from light. After permeabilization with intracellular Staining Permeabilization Wash Buffer Biolegend, USA), 1.5 µL of anti-Cd206 (PE) antibody was added and incubated for 20 min. Finally, the cells were suspended in PBS and analyzed promptly.

### ELISA

Ccl17 and Ccl22, in BALF were analyzed by ELISA (BOSTER Biological Technology, China). ELISA was performed according to the manufacturer’s instructions. All samples and standards were measured in duplicate.

### Immunoprecipitation

NHK 293T cell proteins were extracted using RIPA lysis buffer (Sigma-Aldrich). An appropriate dilution of either anti-FLAG mAb (Proteintech, 1:550 dilution) or anti-GFP mAb (Proteintech, 1:50 dilution) was added to a centrifuge tube coated with Protein G. The antibody-bead mixture was incubated at 4°C for 4 hours using a tube rotator. Then, 50 μg of cell lysates were added to the mixture, and the lysate-bead/antibody conjugate mixtures were incubated overnight at 4°C. The mixtures were subsequently resuspended in 5×SDS loading buffer, boiled for 5 minutes, and analyzed by Western blotting.

### Mass spectrometry analysis

We transfected HEK-293T cells with CDH26-FLAG overexpression plasmid. The samples were obtained through co-immunoprecipitation experiments using the FLAG antibody. Beads samples were incubated in the reaction buffer (1% SDC/100 mM Tris-HCl, pH 8.5/10 mM TCEP/40 mM CAA) at 95 °C for 10 min for protein denaturation, cysteine reduction and alkylation. The eluates were diluted with equal volume of H_2_O and subjected to trypsin digestion overnight by adding trypsin at a ratio of 1:50 (enzyme: protein, w/w) for overnight digestion at 37 °C. The next day, TFA was used to bring the pH down to 6.0 to end the digestion. After centrifugation (12000×g, 15 min), The peptide was purified using self-made SDB desalting columns. The eluate was vacuum dried and stored at -20 °C for later use. Mass spectrometry (MS) data acquisition was carried out on a Q Exactive HF mass spectrometer coupled with UltiMate 3000 RSLCnano system. Peptides were loaded through auto-sampler and separated in a C18 analytical column (75μm × 25cm, C18, 1.9μm, 100Å). Mobile phase A (0.1% formic acid) and mobile phase B (80% ACN, 0.1% formic acid) were used to establish the separation gradient. A constant flow rate was set at 300 nL/min. For DDA mode analysis, each scan cycle is consisted of one full-scan mass spectrum (R = 60 K, AGC = 3e6, max IT = 20 ms, scan range = 350–1800 m/z) followed by 20 MS/MS events (R = 15 K, AGC = 2e5, max IT = 50 ms). HCD collision energy was set to 28. Isolation window for precursor selection was set to 1.6 Da. Former target ion exclusion was set for 30 s. MS raw data were analyzed with MaxQuant (V1.6.6) using the Andromeda database search algorithm. Spectra files were searched against Human database (2022-03-29,20377 entries) using the following parameters: LFQ mode was checked for quantification; Variable modifications, Oxidation (M), Acetyl (Protein N-term) & Deamidation (NQ); Fixed modifications, Carbamidomethyl (C); Digestion, Trypsin/P; The MS1 match tolerance was set as 20 ppm for the first search and 4.5 ppm for the main search; the MS2 tolerance was set as 20 ppm. Search results were filtered with 1% FDR at both protein and peptide levels. Proteins denoted as decoy hits, contaminants, or only identified by sites were removed, the remaining identifications were used for further quantification analysis. Proteins with a fold change > 4 between bait IP and control were screened out as interactors of the bait protein.

### Protein-protein docking

Using the predicted structures of CDH26 (uniProt ID: Q8IXH8) and STUB1 (uniProt ID: Q9UNE7) from the AlphaFold protein structure database (https://alphafold.ebi.ac.uk/), we performed protein-protein docking experiments. Since CDH26 is a transmembrane protein, we extracted the intracellular sequence of CDH26 (636–832) for the docking experiments (http://zdock.umassmed.edu/). We selected the top-scoring docking conformation and identified the binding sites using PDBePISA (https://www.ebi.ac.uk/msd-srv/prot_int/cgi-bin/piserver).

### Preparation of *Cdh26* siRNA encapsulated lipid nanoparticles

*Cdh26* siRNA encapsulated lipid nanoparticles were prepared by Cynbio (Shanghai, China). The preparation method is as follows: *Cdh26* siRNA was dissolved in RNase-Free water. For the aqueous phase, a siRNA working solution was prepared using 100 mM sodium acetate (pH 4.0), with a concentration of 0.35 mg/mL. For the organic phase, a lipid mixture was prepared in anhydrous ethanol at a concentration of 25 mM, with the following composition: DLin-MC3-DMA: DSPC: Cholesterol: DMPE-PEG2000 = 50:10:38.5:1.5. The encapsulation of *Cdh26* siRNA into lipids was performed using the PNI Ignite instrument with the Ignite microfluidic chip, at a total flow rate of 12 mL/min and a water-to-organic ratio of 3:1. After encapsulation, the nanoparticles were diluted with 10 times the volume of DPBS (calcium and magnesium free). The diluted mixture was then centrifuged at 4°C and 3000 g for 30 minutes using a 100 kDa ultrafiltration tube, until the volume before dilution was reached. The buffer was replaced with DPBS three times, achieving a displacement rate of 99%. A sucrose solution of the corresponding concentration was prepared using DPBS, and the *Cdh26* siRNA encapsulated lipid nanoparticles were diluted with a 5% sucrose solution. The encapsulation efficiency, particle size, PDI, and zeta potential were measured. Finally, the *Cdh26* siRNA encapsulated lipid nanoparticles were stored at -80°C.

### Statistical analysis

Data analysis was conducted using Prism version 8 (GraphPad Software). For normally distributed data, we calculated the means ± standard deviation (SD) and performed parametric tests such as Student’s t-test or one-way ANOVA followed by Tukey’s multiple comparison test for group comparisons. For non-normally distributed data, we calculated medians with interquartile ranges and utilized non-parametric tests, such as the Mann-Whitney test. Spearman correlation coefficient was used to assess correlation. Statistical significance was defined as *p* < 0.05.

## Supplementary Figures

**Figure S1.**
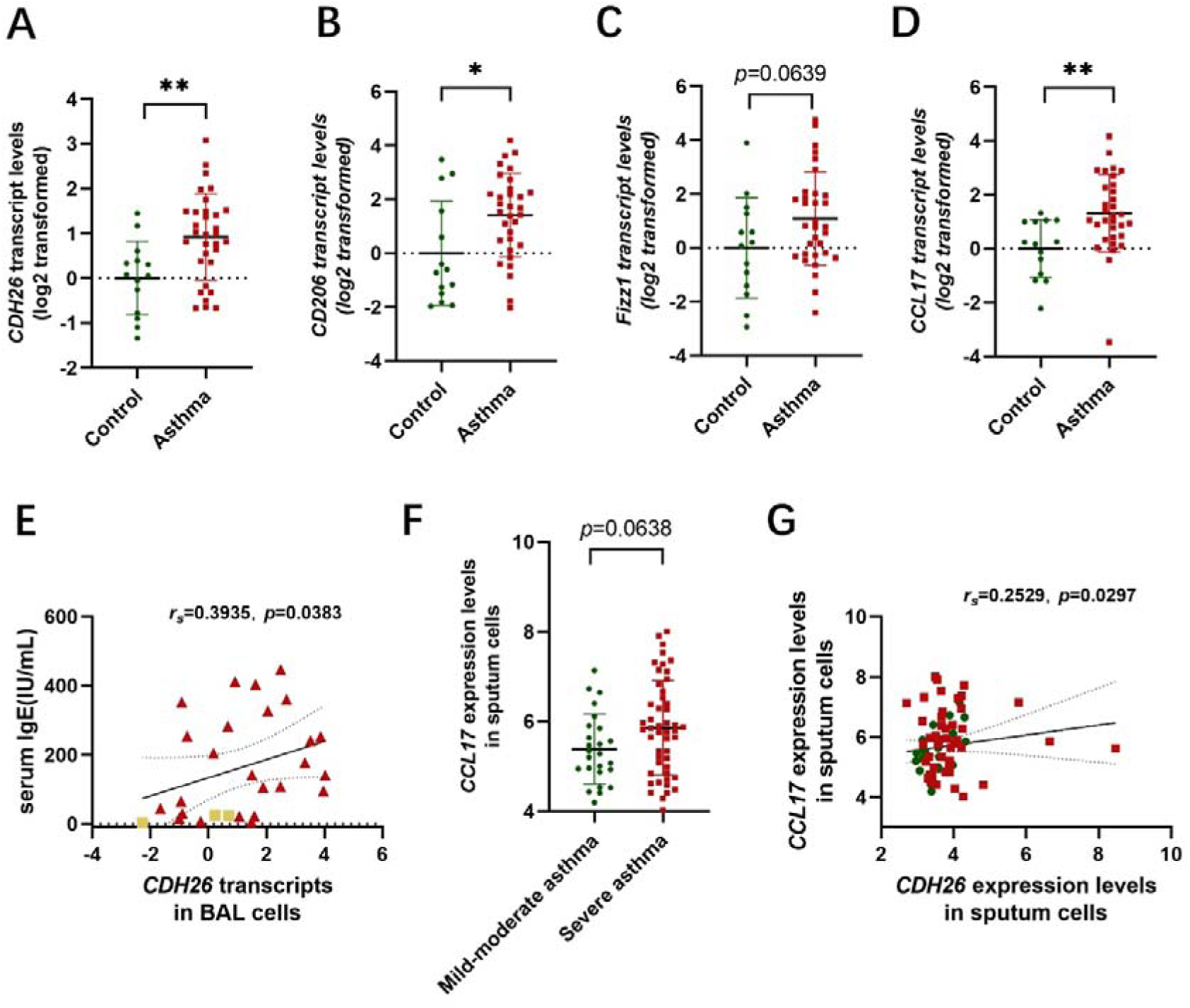
The expression of *CDH26* and M2 macrophage markers are upregulated in induced sputum cells of asthma patients. A-D) *CDH26*, *CD206*, *FIZZ1* and *CCL17* transcript levels in induced sputum cells from asthma patients (n=32) and control subjects (n=14) were measured by quantitative PCR. The transcript levels were expressed as log2 transformed and relative to the mean value for control group. E) Spearman’s correlation assays between *CDH26* transcripts in BAL cells and serum IgE of asthma patients. F) The expression of CCL17 in induced sputum array data (GSE76262). G) Spearman’s correlation assays between *CDH26* and *CCL17* expression levels in asthma patients in induced sputum array data. Data are mean ± SD. \**p*<0.05; \*\**p*<0.01. Abbreviations: CDH26, cadherin-26; CD206, mannose receptor C-Type 1; Fizz1, found in inflammatory zone 1; CCL17, C-C motif chemokine ligand 17.

**Figure S2.**
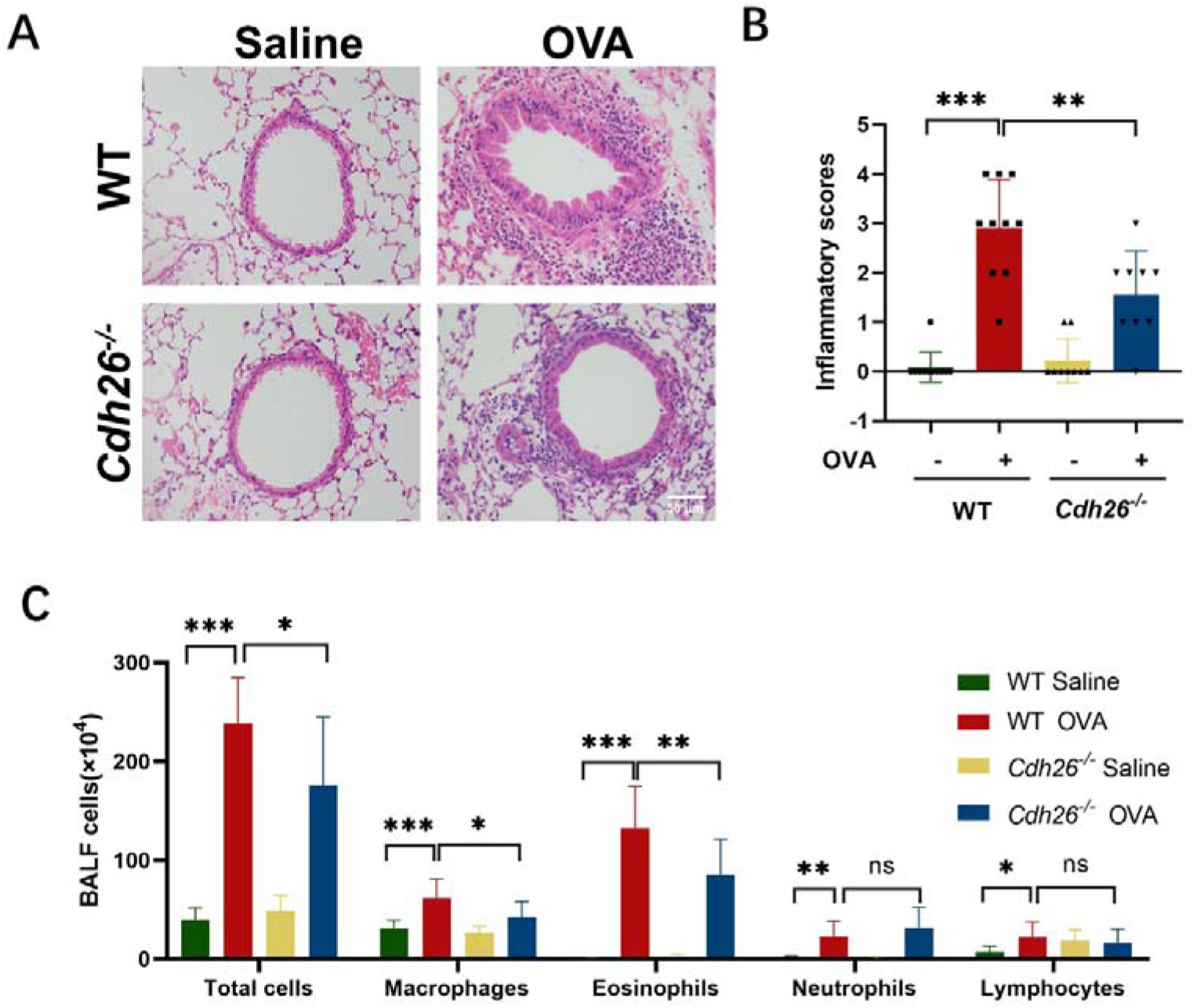
*Cdh26* deficiency suppresses airway inflammation in a mouse model of allergic airway inflammation. A) Representative images of H&E staining in mouse lung sections (scale bar = 50 μm). B) The inflammatory score was graded in four random fields for each lung section at 200× magnification. C) Cell counts for macrophages, eosinophils, lymphocytes and neutrophils in BALF. n=8-10 mice per group. Data are mean ± SD. \**p*<0.05; \*\**p*<0.01; \*\*\**p*<0.001. Abbreviations: H&E staining, hematoxylin-eosin staining; PAS staining, periodic acid-schiff staining; BALF, bronchoalveolar lavage fluid.

**Figure S3.**
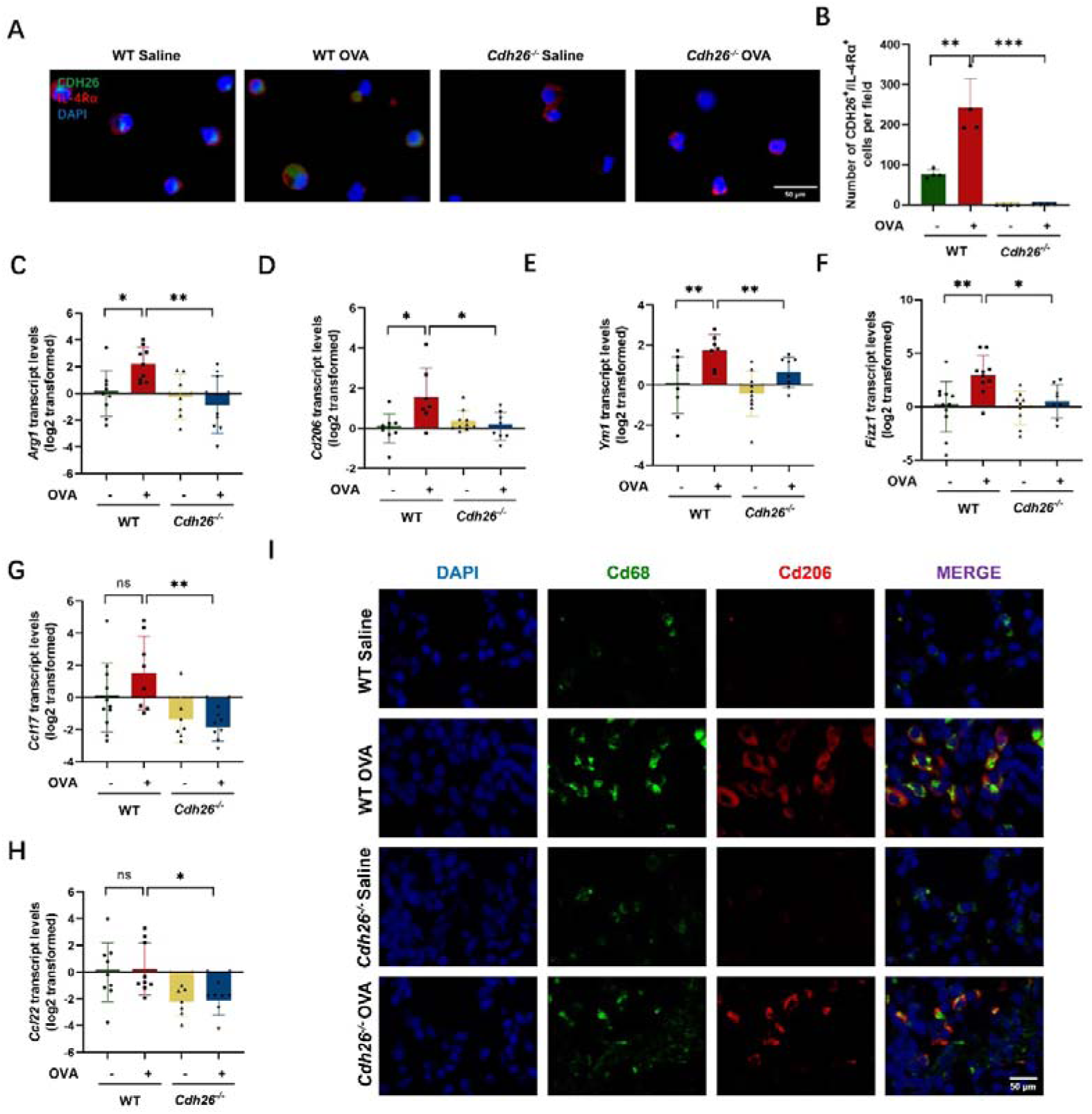
*Cdh26* deficiency suppresses the macrophage alternative activation in a mouse model of allergic airway inflammation. A) Representative images for co-immunofluorescence staining of Cdh26 (green) and Il-4Rα (red) in BALF from mouse (scale bar = 50 μm). Nuclei was stained with DAPI (blue). B) The numbers of Cdh26^+^/ Il-4Rα^+^ cells in BALF were performed using ImageJ. C-H) The mRNA levels of *Arg1*, *Cd206*, *Fizz1*, *Ym1*, *Ccl22* and *Ccl17* in mouse lungs measured by quantitative PCR. The transcript levels were expressed as log2 transformed and relative to the mean value for control group. I) Representative images for co-immunofluorescence staining of Cd68 (green) and Cd206 (red) in BALF from mouse (scale bar = 50 μm). n=8-10 mice per group. Data are mean ± SD. \**p*<0.05; \*\**p*<0.01; \*\*\**p*<0.001. Abbreviations: Cd68, Cd68 molecule; Cd206, mannose receptor C-type 1; Arg1, arginase-1; YM1, also named as Chil3, chitinase-like protein 3; Fizz1, found in inflammatory zone 1; Ccl17, C-C motif chemokine ligand 17; Ccl22, C-C motif chemokine ligand 22; Il-4Rα, lnterleukin-4 receptor subunit alpha.

**Figure S4.**
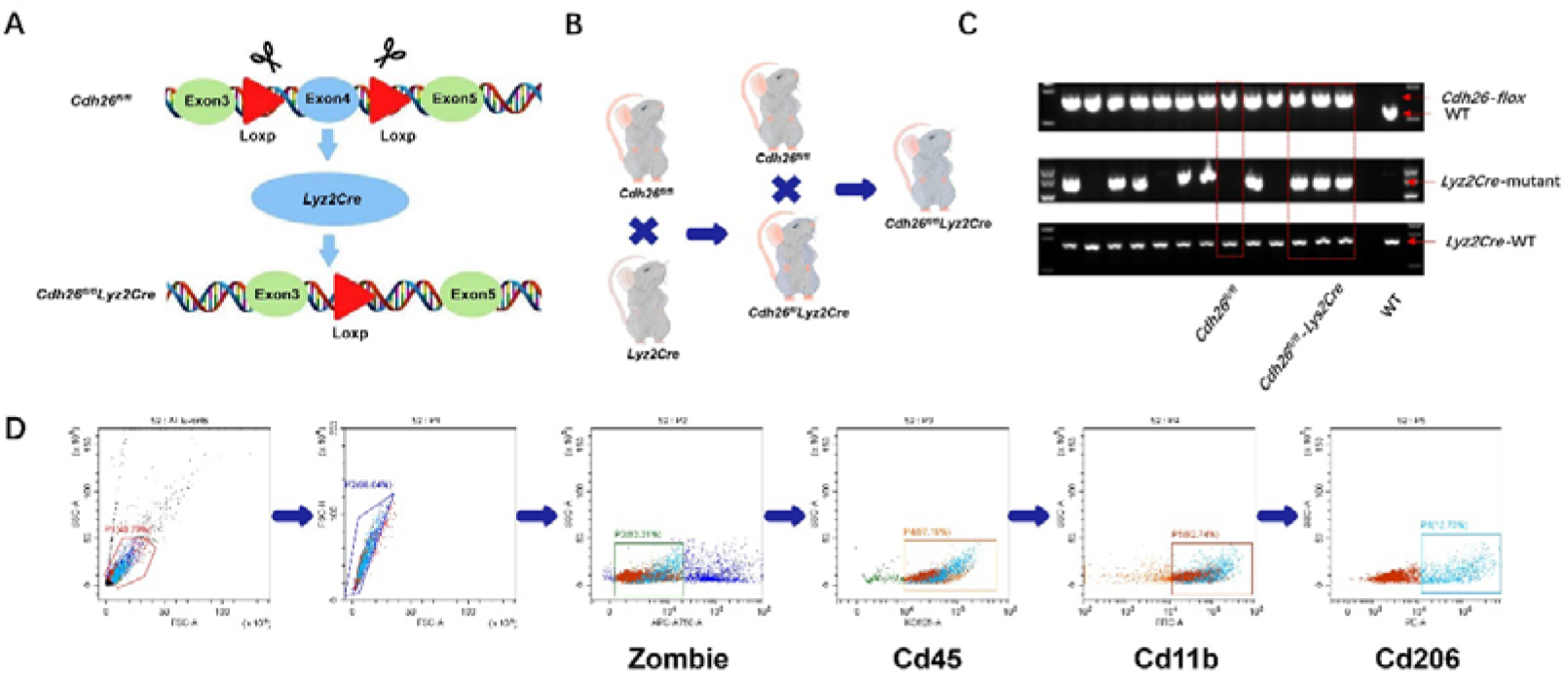
Construction and breeding strategies for *Cdh26^fl/fl^ Lyz2Cre* mice. A) Construction strategies for *Cdh26^fl/fl^ Lyz2Cre* mice. B) Breeding strategies for *Cdh26^fl/fl^ Lyz2Cre* mice. C) Genetic identification *Cdh26^fl/fl^ Lyz2Cre* mice. D) Gating strategies for flow cytometry. Abbreviations: Cd45, Cd45 antigen; Cd11b, Cd11 antigen-like family member b; Cd206, mannose receptor C-type 1.

**Figure S5.**
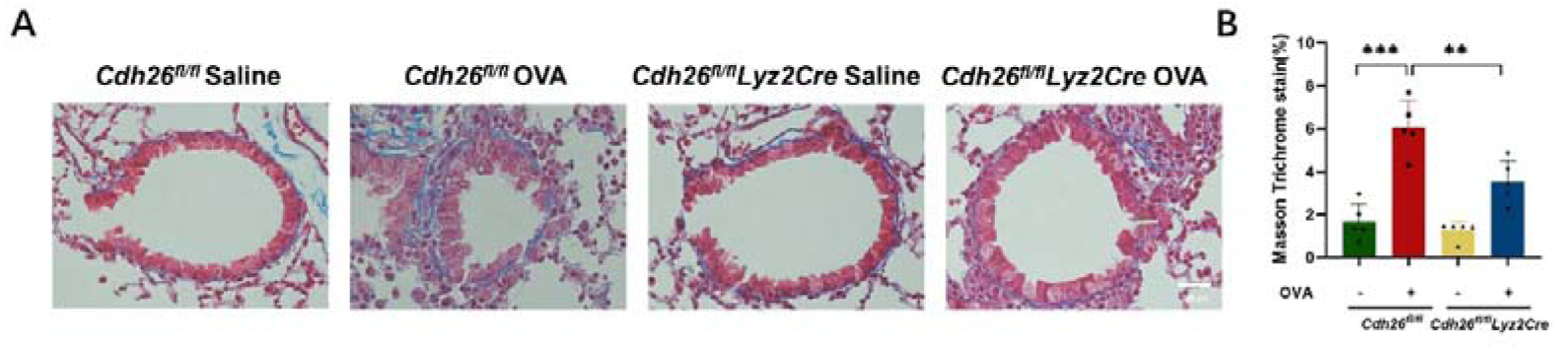
*Cdh26* deficiency inhibits airway remodeling in a mouse model of allergic airway inflammation. A) Representative images of masson staining in mouse lung sections (scale bar = 50 μm). B) The area of masson trichrome stain were measured by ImageJ software. Data are mean ± SD. \*\**p*<0.01; \*\*\**p*<0.001.

**Figure S6.**
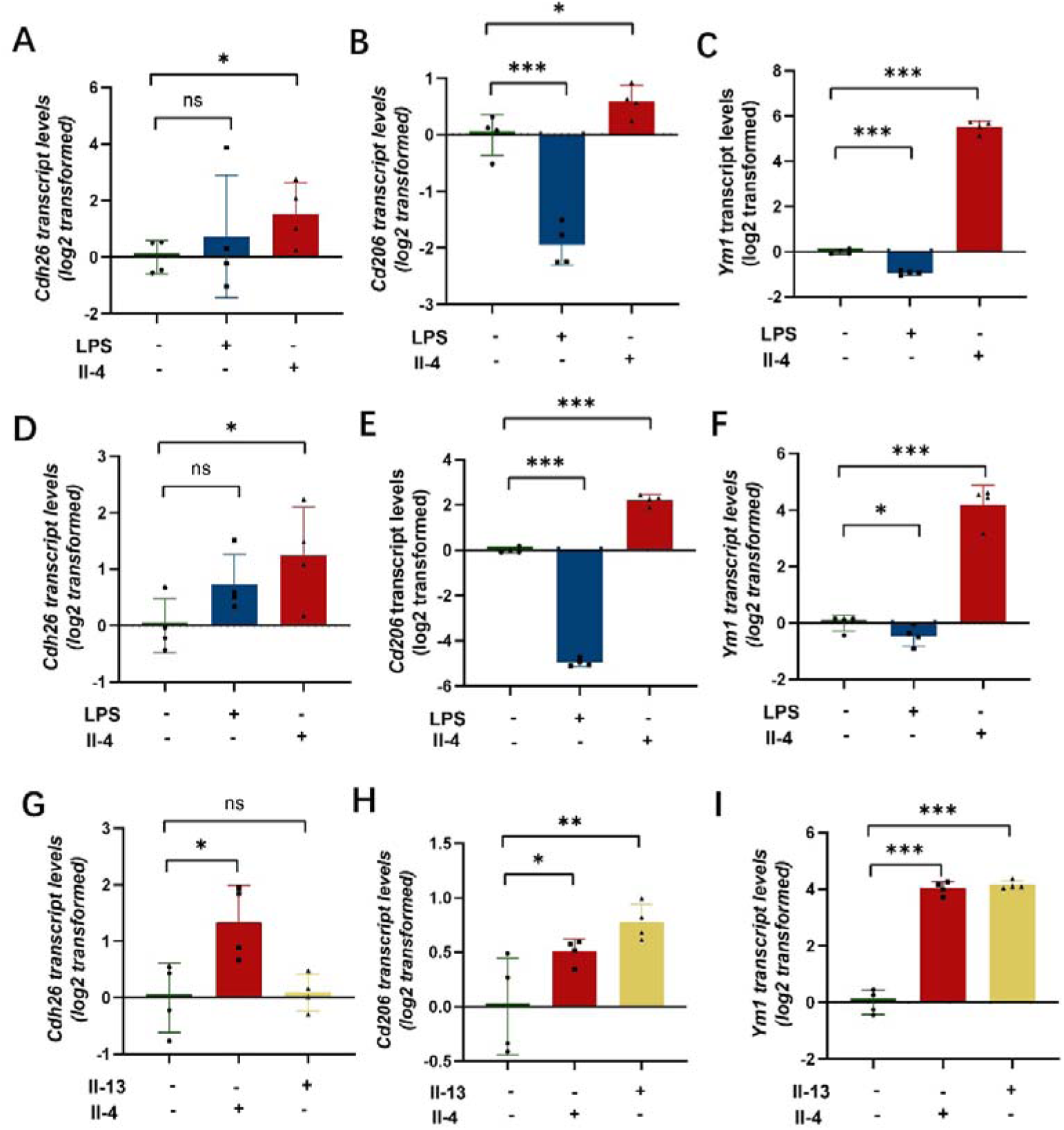
IL-4 upregulates CDH26 in primary cultures of mouse bone marrow-derived macrophages and mouse lung macrophages. A-C) Primary culture of mouse bone marrow-derived macrophages (BMDMs) were stimulated with Il-4 or LPS for 48h. The mRNA levels of *Cdh26*, *Cd206* and *Ym1* were determined by quantitative PCR. D-E) Primary culture of mouse pulmonary macrophages were stimulated with Il-4 or LPS for 48h. The mRNA levels of *Cdh26*, *Cd206* and *Ym1* were determined by quantitative PCR. G-I) Primary culture of BMDMs were stimulated with Il-4 or Il-13 for 48h. The mRNA levels of *Cdh26*, *Cd206* and *Ym1* were determined by quantitative PCR. The transcript levels were expressed as log2 transformed and relative to the mean value for control group. Data are mean ± SD. \**p*<0.05; \*\**p*<0.01; \*\*\**p*<0.001. Abbreviations: Cdh26, cadherin-26; Cd206, mannose receptor C-type 1; YM1, also named as Chil3, chitinase-like protein 3; LPS, lipopolysaccharides.

**Figure S7.**
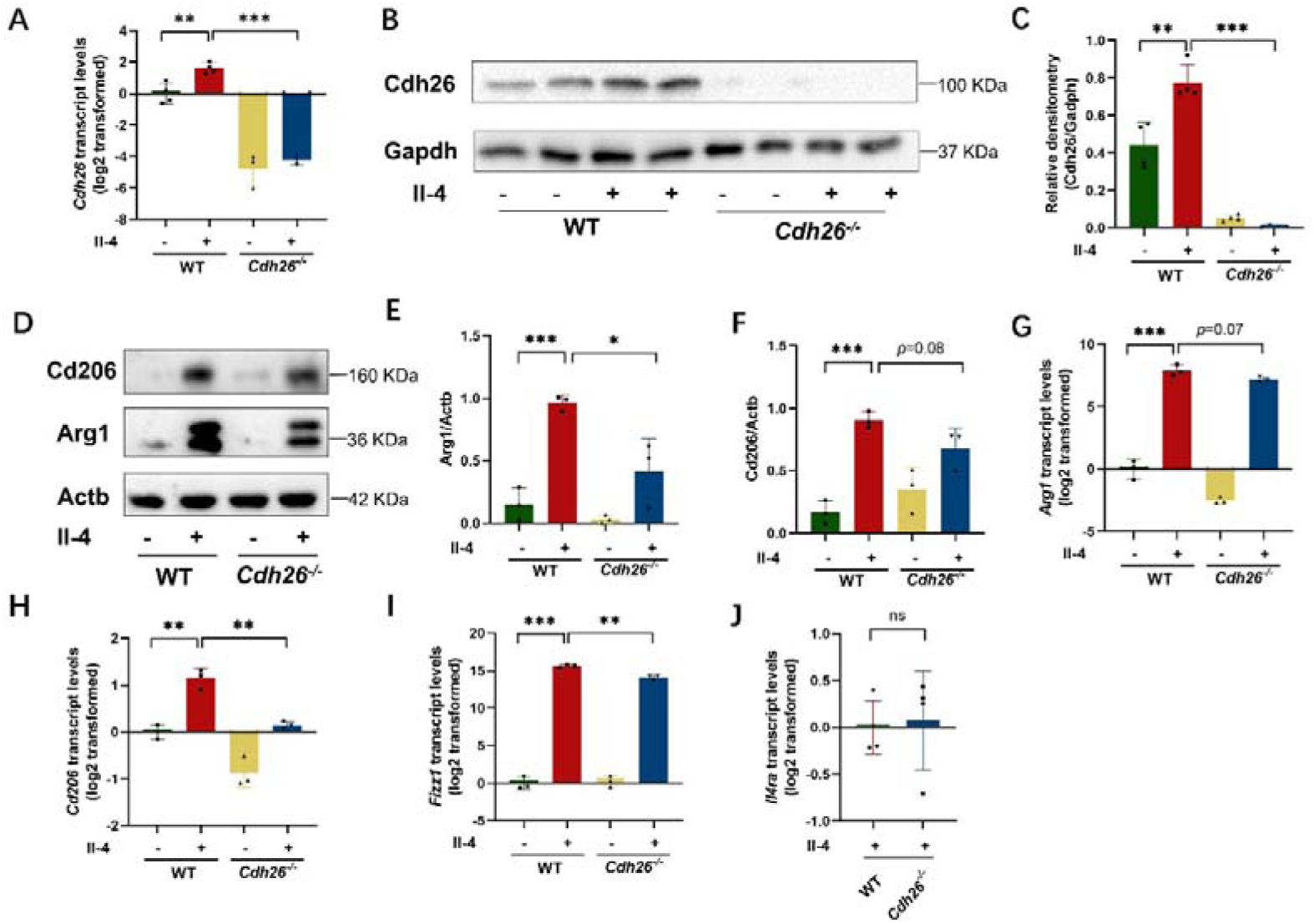
*Cdh26* deficiency suppresses M2 polarization of cultured mouse bone marrow-derived macrophages in. A) Primary culture of mouse bone marrow-derived macrophages (BMDMs) were stimulated with Il-4 for 48h. The mRNA levels of *Cdh26* were determined by quantitative PCR. The transcript levels were expressed as log2 transformed and relative to the mean value for control group. B-C) The protein levels of Cdh26 in BMDM cells after exposure to Il-4 for 48h were determined by western blotting. Densitometry assay was performed using ImageJ, and Cdh26 protein levels were indexed to Gapdh. D-F). The protein levels of Arg1, Cd206 in BMDM cells after exposure to Il-4 for 48h were determined by western blotting. Densitometry assay was performed using ImageJ, and Arg1, Cd206 protein levels were indexed to Actb. G-J) The mRNA levels of *Arg1*, *Cd206*, *Fizz1* and *Il4ra* were determined by quantitative PCR. The transcript levels were expressed as log2 transformed and relative to the mean value for control group. Data are mean ± SD. \**p*<0.05; \*\**p*<0.01; \*\*\**p*<0.001. Abbreviations: Cdh26, cadherin-26; BMDM, bone marrow derived macrophage; Cd206, mannose receptor C-type 1; Arg1, arginase-1; Fizz1, found in inflammatory zone 1; Actb, actin beta; Gapdh, glyceraldehyde-3-phosphate dehydrogenase.

**Figure S8.**
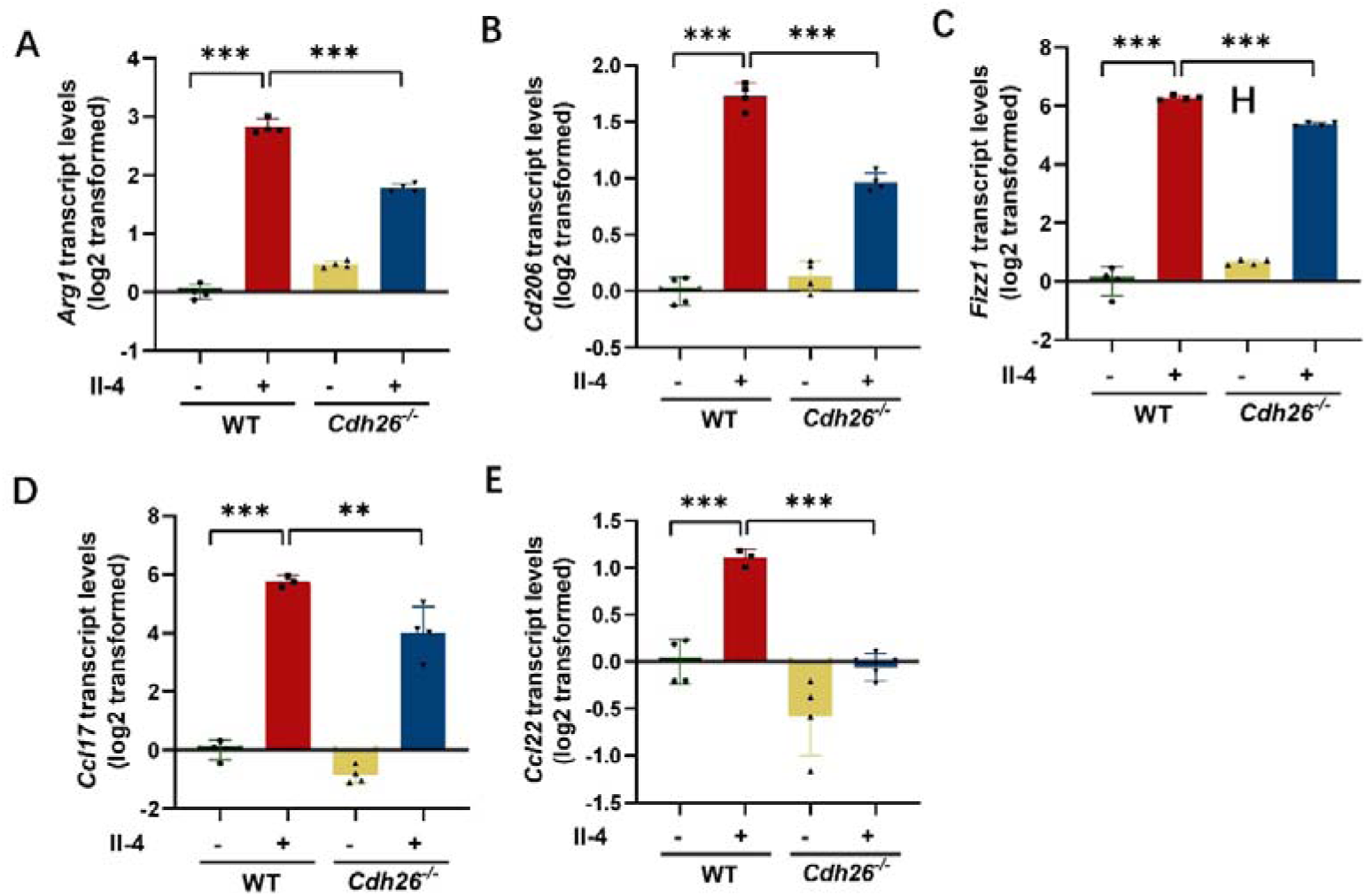
*Cdh26* deficiency suppresses M2 polarization of cultured mouse lung macrophages. A-E) Primary culture of mouse lung macrophages were stimulated with Il-4 for 48h. The mRNA levels of Arg1, Cd206 and Fizz1, Ccl17 and Ccl22 were determined by quantitative PCR. The transcript levels were expressed as log2 transformed and relative to the mean value for control group. Data are mean ± SD. \**p*<0.05; \*\**p*<0.01; \*\*\**p*<0.001. Abbreviations: Arg1, arginase-1; Cd206, mannose receptor C-type 1; Fizz1, found in inflammatory zone 1; Ccl17, C-C motif chemokine ligand 17; Ccl22, C-C motif chemokine ligand 22, Actb, actin beta.

**Figure S9.**
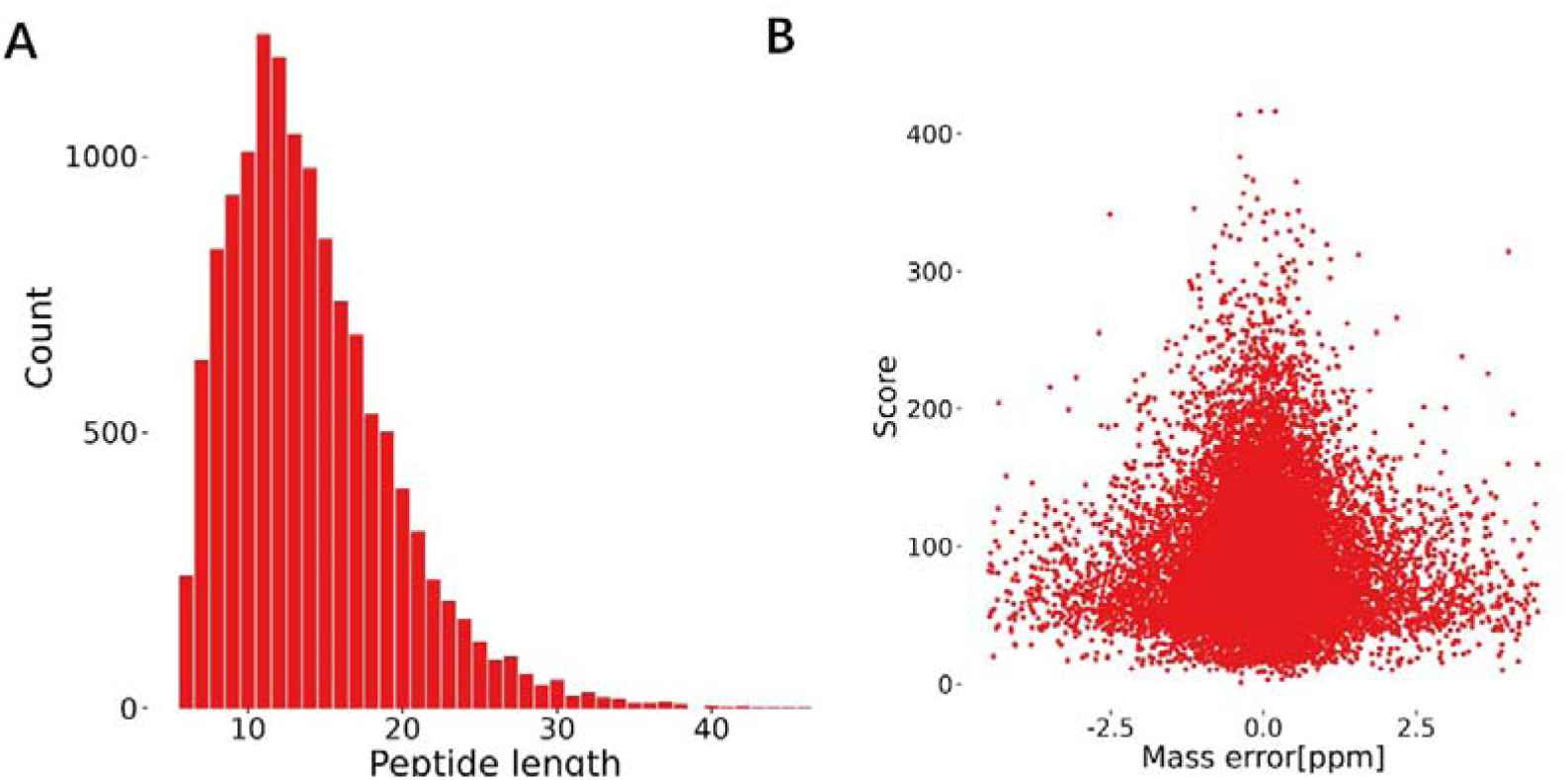
The quality control for mass spectrometry analysis. A) The peptide length distribution of mass spectrometry analysis. B) The mass deviation distribution plot of mass spectrometry analysis.

**Figure S10.**
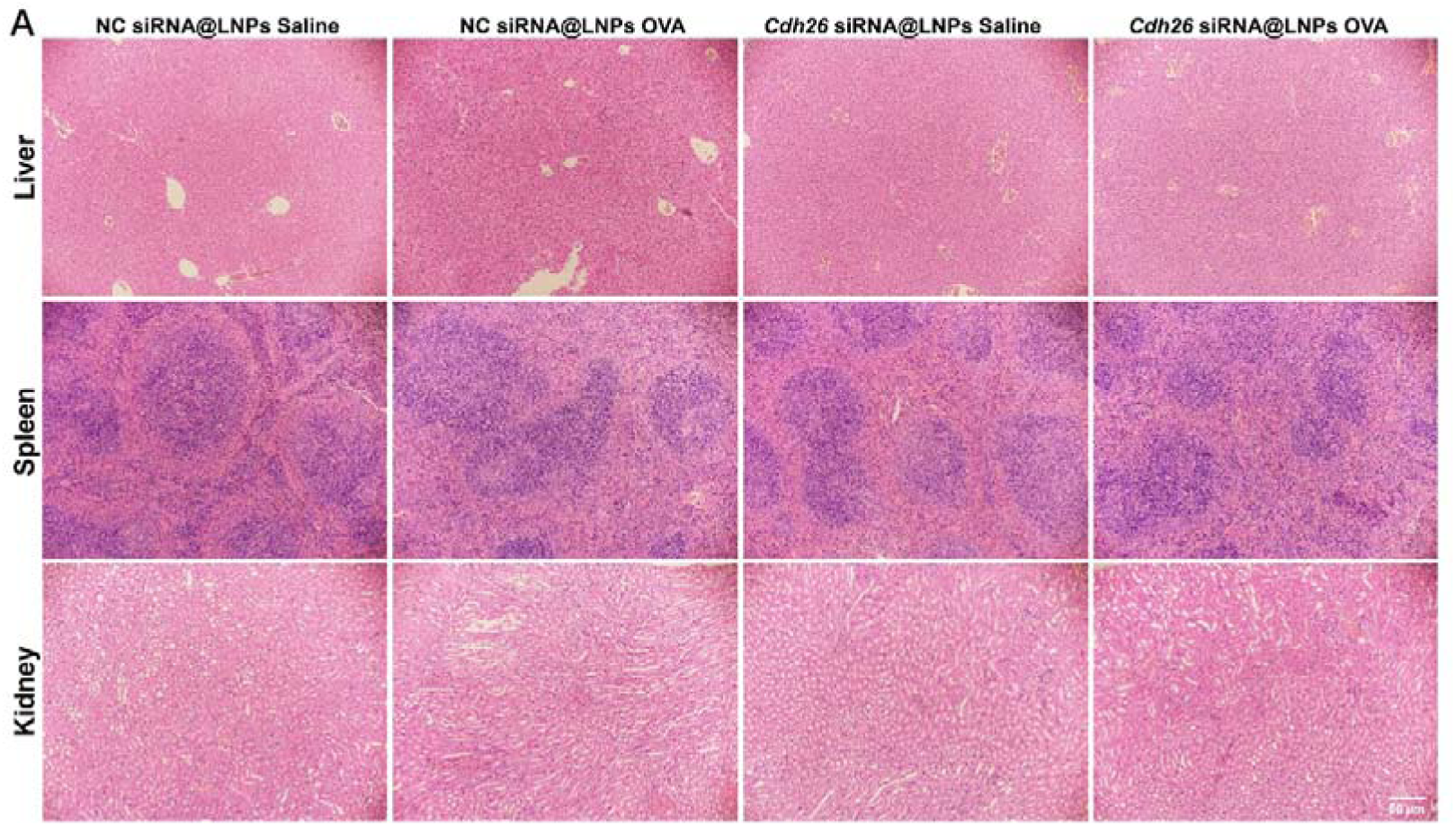
H&E staining of liver, spleen, and kidney of the mice treated with siRNA encapsulated lipid nanoparticles. WT mice were treated with NC siRNA@LNPs + Saline, NC siRNA@LNPs + OVA, *Cdh26* siRNA@LNPs + Saline or *Cdh26* siRNA@LNPs + OVA, respectively. Representative images for H&E staining of liver, spleen, and kidney. n=8-10 mice per group.

**Figure S11.**
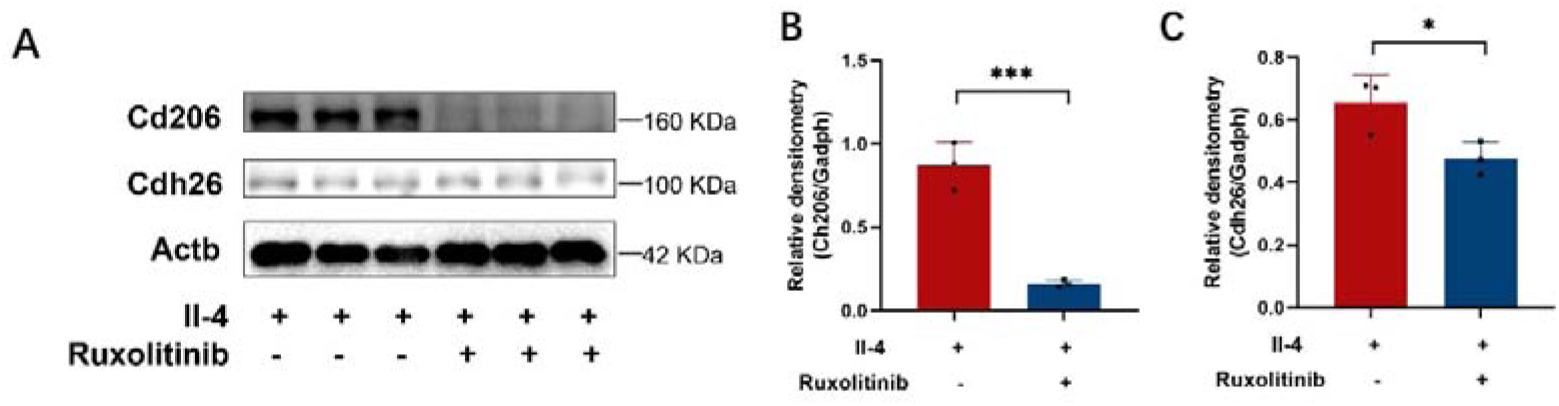
JAK1 inhibitor inhibits CDH26 expression in IL-4-stimulated mouse bone marrow-derived macrophages. A-C) Primary culture of mouse BMDM cells were pretreated with ruxolitinib for 2h and stimulated with Il-4 for 48h. The protein levels of Cdh26, Cd206 were determined by western blotting. Densitometry assay was performed using ImageJ, and Cdh26, Cd206 protein levels were indexed to Actb. Data are mean ± SD. \**p*<0.05; \*\*\**p*<0.001. Abbreviations: Cdh26, cadherin-26; Cd206, mannose receptor C-type 1; Actb, actin beta.

**Table S1.**
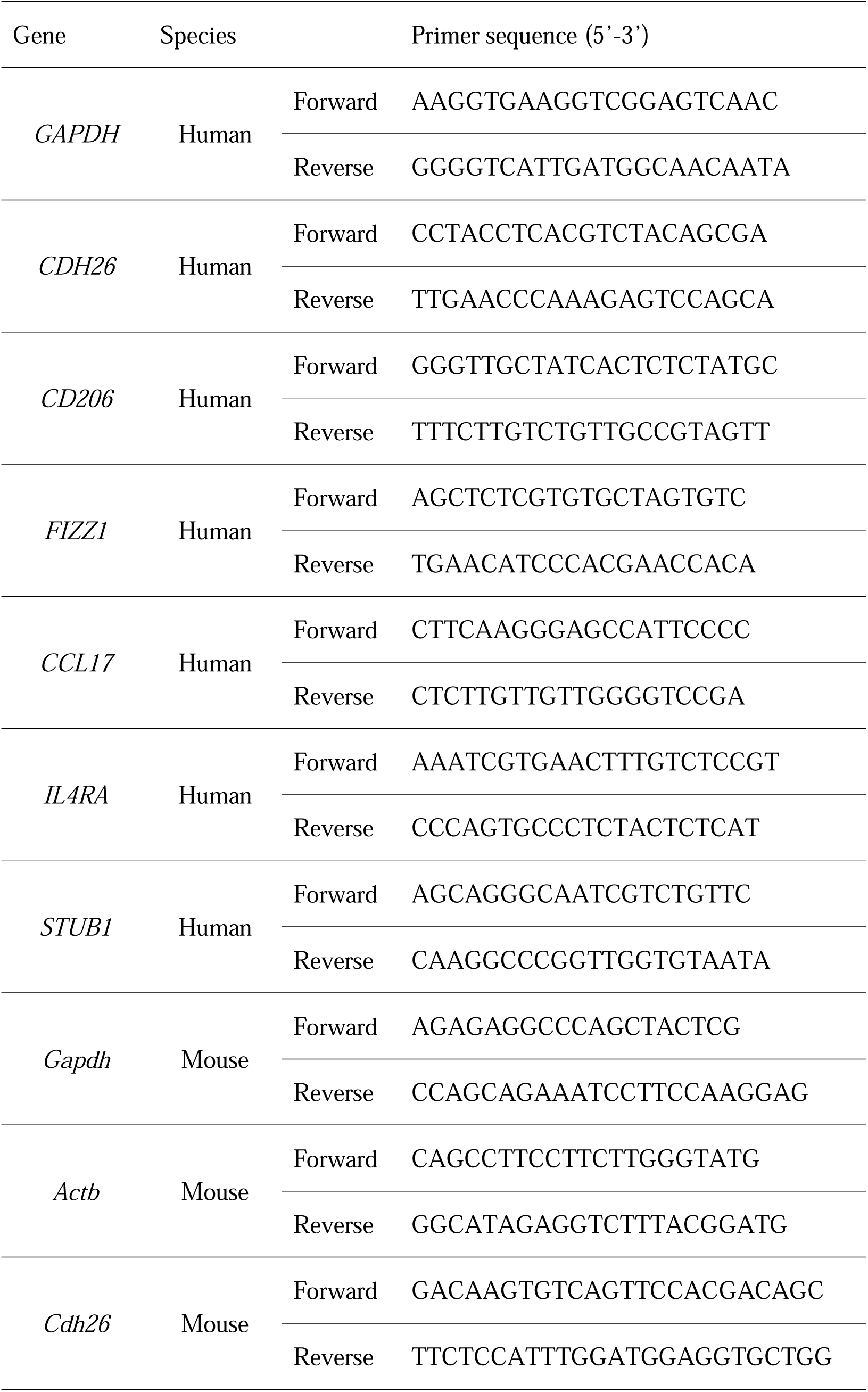
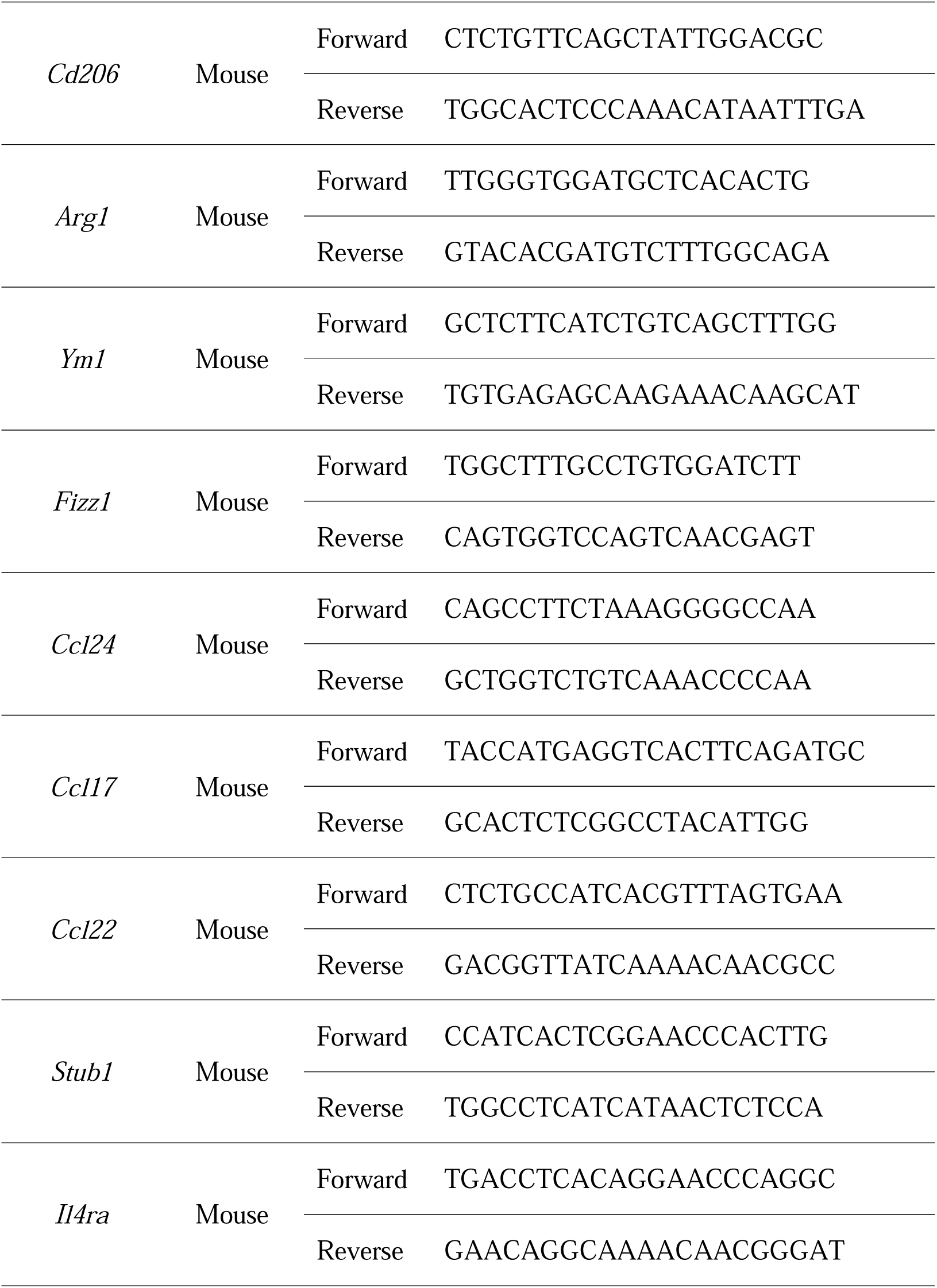
Primers for quantitative PCR

**Table S2.**
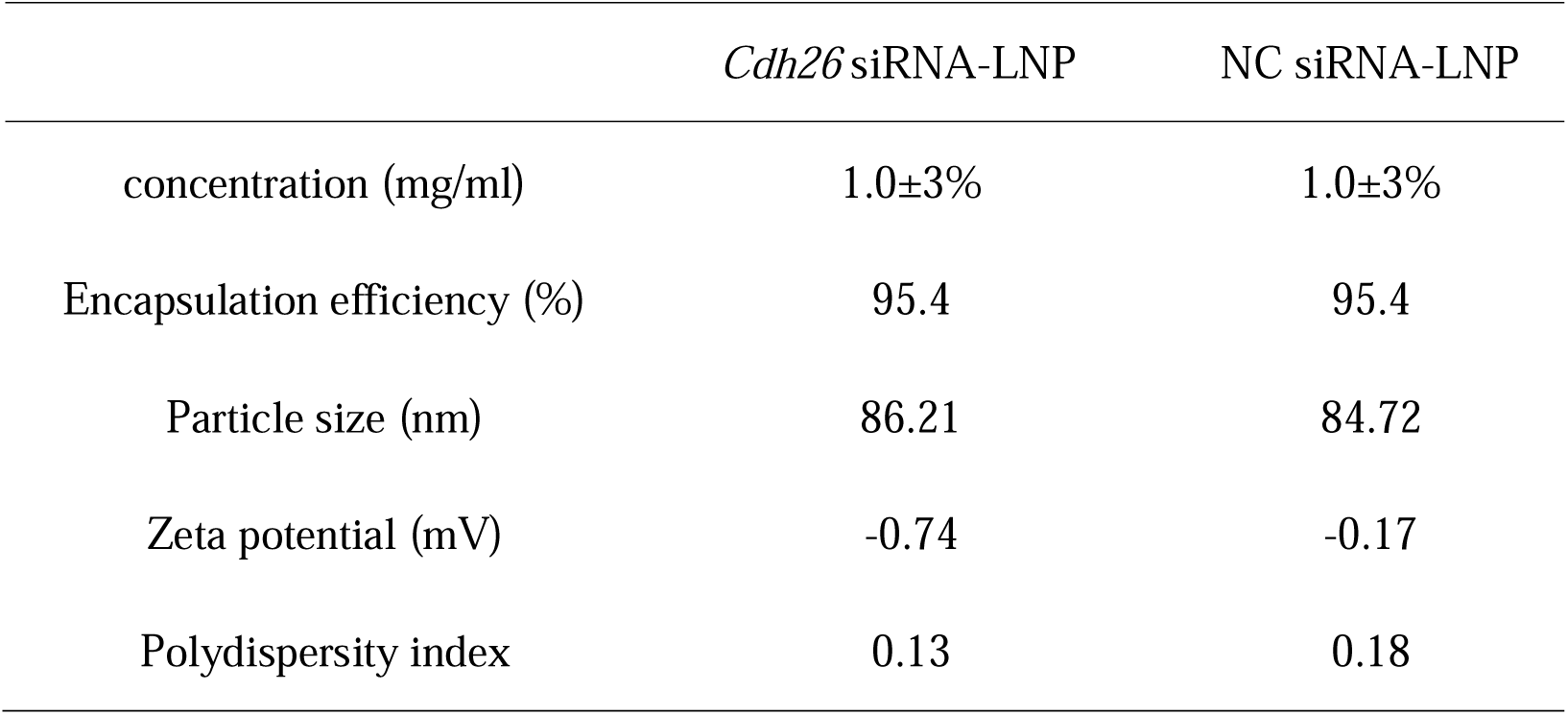
The characterization of siRNA encapsulated lipid nanoparticles.

|  | *Cdh26* siRNA-LNP | NC siRNA-LNP |
| --- | --- | --- |
| concentration (mg/ml) | 1.0±3% | 1.0±3% |
| Encapsulation efficiency (%) | 95.4 | 95.4 |
| Particle size (nm) | 86.21 | 84.72 |
| Zeta potential (mV) | -0.74 | -0.17 |
| Polydispersity index | 0.13 | 0.18 |

## Notes

### Competing Interest Statement

The authors have declared no competing interest.

### Author Declarations

The study was approved by the ethics committee of Tongji Hospital, Tongji Medical College, Huazhong University of Science and Technology.

### Summary of Updates

Figure 1，6，7，8 revised.Author affiliations updated

